# Early Diagnosis and Prognostic Prediction of Colorectal Cancer through Plasma Methylation Regions

**DOI:** 10.1101/2024.11.28.24317652

**Authors:** Lingqin Zhu, Lang Yang, Fangli Men, Jianwei Yu, Shuyang Sun, Chenguang Li, Xianzong Ma, Junfeng Xu, Yangjie Li, Ju Tian, Xin Wang, Hui Xie, Qian Kang, Linghui Duan, Xiang Yi, Wei Guo, Xueqing Gong, Ni Guo, Youyong Lu, Joseph Leung, Yuqi He, Jianqiu Sheng

## Abstract

Cell-free DNA (cfDNA) methylation is a valuable biomarker in various cancers including colorectal cancer (CRC), but marker for both early diagnosis and prognostic prediction remain a critical unmet need. Here, we report the development of a 27-DMR (differentially methylated regions) plasma panel with dual diagnostic and prognostic functions. We first identified CRC-specific methylation features from 119 tissue samples and 161 plasma samples. Machine learning algorithms were then applied to develop diagnosis and prognosis models using the plasma samples in training cohort. In the tissue external validation cohort (GSE48684), the cfDNA methylation diagnosis model conducted with the panel, achieved an area under the curve (AUC) of 0.983, and for the plasma cfDNA model in the external validation cohort, the sensitivities for NAA, AA and CRC 0 - II are 48.4%, 52.2% and 66.7% respectively, with a specificity of 88%. Beyond its diagnostic capabilities, the methylation panel was also a powerful predictor of distant metastasis (AUC = 0.955) and patient prognosis (AUC = 0.867). Using normal samples as control, the changes in methylation score in both tissue and plasma were consistent across different lesions, although the degree of alterations varied with severity. The methylation scores vary between paired tissue and blood samples, suggesting distinct mechanisms of migration from tumor tissue to blood for the 27 DMRs. Together, our cfDNA methylation models based on 27 DMRs can identify different stages of CRC and predict metastasis and prognosis, ultimately enabling early intervention and risk stratification for CRC patients.

## Introduction

Colorectal cancer (CRC) is the third most prevalent malignancy and the fourth leading cause of cancer-related mortality worldwide, accounting for 9.4% of all cancer deaths (^1^). When patients were diagnosed with CRC, over half of their family (62.9%) faced financial burdens (^2, 3^). The survival rates of individuals diagnosed with advanced CRC (Stage III or IV) witness a significant decrement (^4, 5^), underscoring early detection and removal of precancerous lesions remain the most effective strategy to prevent CRC-associated mortality (^6^). Although colonoscopy is the gold standard for CRC detection, its invasiveness, suboptimal patient compliance and risk of complications including intestinal perforation limit its widespread effectiveness (^7^). Other non-invasive tests, such as guaiac-based fecal occult blood test (gFOBT), fecal immunochemical test (FIT), carcino-embryonic antigen (CEA), and multi-target stool DNA (mt-sDNA) testing, are constrained by insufficient sensitivity, particularly for early-stage lesions (^8, 9^).

Recent research demonstrated a profound correlation between CRC and the development of genetic and epigenetic alterations. During early stage of tumorigenesis, epigenetic modifications surpass the frequency of gene mutations, indicating their potential as diagnostic biomarkers in screening for colon polyps and cancer (^10^). Circulating tumor DNA (ctDNA), a component of cell-free DNA (cfDNA) released from apoptotic and necrotic tumor cells, carries cancer-specific epigenetic alterations (^11^). Consequently, cfDNA methylation emerges as a promising biomarker for cancer screening due to its early onset and high signal density(^12^).

While the utility of cfDNA methylation in early diagnosis, surveillance, molecular subtyping and prognostic prediction of CRC has been proven, current approaches have limitations (^13^). For instance, a diagnostic panel has shown excellent performance but was developed using inconsistent sample types (e.g., CRC tissue vs. normal blood leukocyte methylation data), introducing potential bias (^14^). Another study addressed the potential of cfDNA methylation in patient risk stratification, but required blood sampling every three months, which can hamper patient compliance and clinical feasibility (^15^).

In this study, we aimed to develop and validate a plasma-based biomarker panel, initially identified from CRC tissues, for the comprehensive diagnosis, metastatic assessment, and prognostic prediction of CRC. We performed a meticulous, multi-stage screening process starting with CRC and normal tissues, followed by refinement using plasma samples. Crucially, by utilizing a substantial cohort of paired tissue and plasma samples from the same patients, we minimized potential confounding biases. The resulting 27 differential methylated regions (DMRs) panel was designed to, from a single blood test, ascertain the presence of CRC, distinguish lesion stages, identify distant metastasis, and stratify patients by risk. Such a multi-functional tool holds significant promise for guiding the clinical management of CRC, from early detection to personalized treatment strategies.

## Materials and Methods

### Study design and samples

This study was designed to first identify CRC-specific DNA methylation markers and then to develop and validate models for CRC diagnosis, metastasis, and prognosis. The marker discovery phase utilized data from TCGA public database and clinically collected tissue and plasma samples. The 450k chip methylation data encompassing colorectal, esophageal, gastric, lung, liver, and breast cancers, and CRC transcriptome sequencing data were downloaded from The Cancer Genome Atlas (TCGA) database. The methylation data of tissue screening cohort was obtained from fifteen pairs of tissue samples sequenced with the Roche Nimble Gen Seq Cap Epi method from the Seventh Medical Center of PLA General Hospital. The 89 tissue samples and 77 plasma samples used for marker screening were collected between June to December 2020 at the Seventh Medical Center of PLA General Hospital and Dongying People’s Hospital. Participants for model establishment and validation were enrolled from December 2020 to July 2022 in the Seventh Medical Center of PLA General Hospital, Dongying People’s Hospital. The paeticipants for external validation cohort were enrolled from July 2022 to December 2022 at the First Hospital of Longyan, Fujian Medical University. An additional public tissue validation cohort (GSE48684) was obtained from the Gene Expression Omnibus (GEO). Detailed patient inclusion and exclusion criteria, as well as methods for DNA extraction, library construction, and targeted bisulfite sequencing, are provided in the Supplementary Materials.

### CRC-specific methylation markers selection

An initial marker pool was generated by integrating TCGA methylation data with our 15 pairs of tissue methylation sequencing results. This informed the design of a targeted panel, which was then screened in the 89 samples (13 normal tissues and 38 paired CRC tissues), identifying 404 DMRs. Then, the candidates were further evaluated in 77 plasma samples from 13 Normal, 15 non-advanced adenoma (NAA), 12 advanced adenoma (AA) and 37 CRC participants. Subsequently, the Least Absolute Shrinkage and Selection Operator (LASSO) regression analysis was used to filter markers, and 27 regions that appeared in more than 50 iterations were chosen as plasma diagnostic markers. The correlation between the methylation levels of the 27 DMRs and the expression of their associated genes was assessed using Spearman’s rank correlation.

### Construction of the CRC diagnosis model

A diagnostic model was developed using a logistic regression algorithm trained on methylation data from the training cohort. The model was built using five-fold cross-validation, optimizing for the minimum binary deviance. Its performance was evaluated by Receiver Operating Characteristic (ROC) curve analysis and the optimal cutoff value for the resulting methylation score was determined using the maximum Youden index. Using the methylation scores derived from the diagnosis model, the CRC staging (AA, CRC 0-II, CRC III-IV) was predicted.

### Construction of the CRC metastasis and prognosis models

A metastasis prediction model was constructed using the methylation score from the diagnostic model combined with patients’ clinical metastasis status. Similarly, a prognostic model was developed by integrating the methylation score with patient survival data. The performance of both models was evaluated using ROC analysis, with optimal cutoffs determined by the maximum Youden index.

## Statistical analysis

All statistical analyses were performed using SPSS 26.0 or R 4.1.3. Two-sided tests were used for p-values, with differences deemed statistically significant at *p* < 0.05. Model performance was assessed by ROC analysis using the “roc” function from the R package pROC, generating the area under the ROC curve (AUC) with 95% confidence interval (CI). Survival analyses were conducted using Kaplan-Meier (KM) curves with the log-rank test and hazard ratios were calculated using Cox proportional-hazards models.

## Results

### Patient characteristics

To delineate DNA methylation biomarkers specific to CRC, 119 tissues and 77 plasma samples (originating from the same patients with tissues) were collected for methylation sequencing analysis, narrowing down to 142 DMRs. Then the 27 DMRs were selected with the sequencing data of 84 plasma samples. Following this, CRC diagnosis, metastasis, and prognosis models were constructed using plasma samples from 84 individuals (51 CRC, 33 Normal). Validation was then carried out in an independent cohort (30 CRC, 37 AA, 14 Normal). The CRC diagnosis model was further tested in an external validation cohort (18 CRC 0-II, 91 NAA, 23 AA, 34 Normal). Detailed information about the study design is illustrated in Figure 1, and comprehensive patient characteristics are summarized in Table s2-s4.

**Figure 1.**
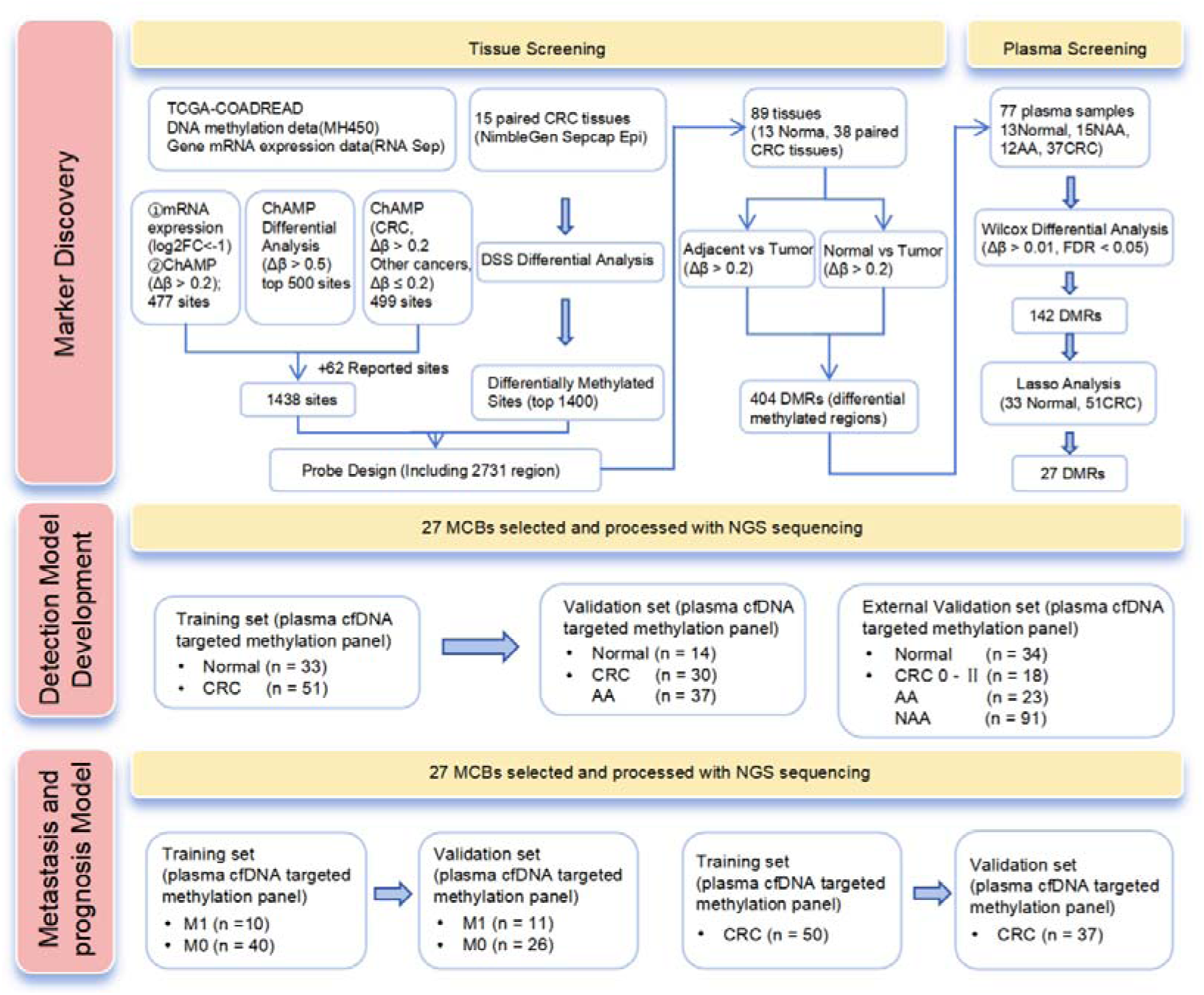
Flow diagram of the study.

### Methylation markers selection

We employed a multi-stage funneling strategy to identify a robust panel of CRC-specific methylation markers, starting from a broad analysis of public data and progressively refining the list through validation in our own tissue and plasma samples. Analyzing the 450k chip methylation data from TCGA, we identified the top 500 differentially methylated CpG sites (DMCs) between CRC and control tissues (Δβ > 0.50, FDR < 0.05), 477 hypermethylated sites (Δβ > 0.20, FDR < 0.05) located in the TSS or First Exon regions of significantly downregulated genes (log2FC < −1, FDR < 0.05), and 499 sites that were hypermethylated in CRC (Δβ > 0.20) but not in five other major cancers (esophageal, gastric, lung, liver, and breast cancer; Δβ ≤ 0.20). The union of these three approaches, supplemented with known CRC-related methylation sites from existing literature, yielded an initial list of 1,438 candidate DMCs (Supplementary Figure 1A). Unsupervised clustering of these sites revealed distinct methylation patterns between normal and tumor tissues in the TCGA cohort (Figure s1B). To validate these findings in our patient population, we performed methylation sequencing (SeqCap Epi) on 15 paired tissue samples (5 advanced adenoma pairs and 10 CRC pairs). From this analysis, we selected the top 1,400 DMCs based on their statistical significance (Δβ > 0.20, FDR < 0.05) (Figure s2A-C). The distribution of the DMCs was predominantly observed in introns and promoters, signifying substantial implications for gene expression and cellular functions (Figure s2D). Merging the 1438 DMCs with the 1400 DMCs for probe design, 404 DMRs were discerned across 89 tissues (13 normal tissues and 38 paired CRC tissues) (Figure s3A). The different methylation patterns between Normal and Tumor were also quite evident (Figure s3B-C). These DMRs correspond to genes that play pivotal roles in biological processes such as cell-cell adhesion, digestive system development, and cAMP and cGMP pathways (Figure s3D). To identify markers detectable non-invasively, we assessed the 404 DMRs in 77 plasma samples (13 Normal, 15 NAA, 12 AA, 37 CRC) and narrowed the list to 142 DMRs that showed significant differential methylation in plasma (Wilcoxon test; Δβ > 0.01, FDR < 0.05) (Figure s3E-F). Finally, to select the most robust and minimal signature for diagnostic modeling, we applied a Least Absolute Shrinkage and Selection Operator (LASSO) regression analysis to the training cohort methylation data. The analysis involved 500 random iterations, and we selected markers that appeared in more than 50 iterations and yielded a final panel of 27 top-performing DMRs for plasma-based CRC detection. Detailed information on these 27 DMRs is provided in Table s5. Correlation analysis using TCGA data confirmed that the methylation levels of the majority of these 27 DMRs were significantly correlated with the expression of their associated genes, underscoring their biological relevance (Figure s4).

### Development and validation of the CRC diagnosis model

Using methylation data from 27 DMRs in tissues, we applied machine learning to construct a diagnosis model for CRC and the AUC reached a high value of 0.994 (Figure 2A). In the external tissue independent validation cohort (GSE48684), the AUC for CRC is 0.983 and for AA is 0.966. (Figure 2B-C). The sensitivities for AA and CRC were 95% and 94%, respectively (Figure 2D-E). Besides, the plasma-based diagnosis model for CRC was constructed with training cohort and validated in an external independent validation cohort, the 27 DMRs exhibited distinctive methylation patterns between CRC and normal individuals (Figure 3A-B). The methylation scores of the model were significantly elevated in the CRC group compared to the normal and AA groups (*P*< 0.001), and they increased with advance tumor staging (Figure 3C). In the training cohort, the ROC curve analysis showed an AUC value of 0.928 [95% confidence interval (CI): 0.874–0.981] (Figure 3D). ROC curve analysis in the validation cohort showed AUC values for AA, stages 0-II, III-IV, and all CRC stages as 0.714 (0.550–0.878), 0.890 (0.749–1.000), 0.967 (0.899–1.000), and 0.929 (0.827–1.000).

**Figure 2.**
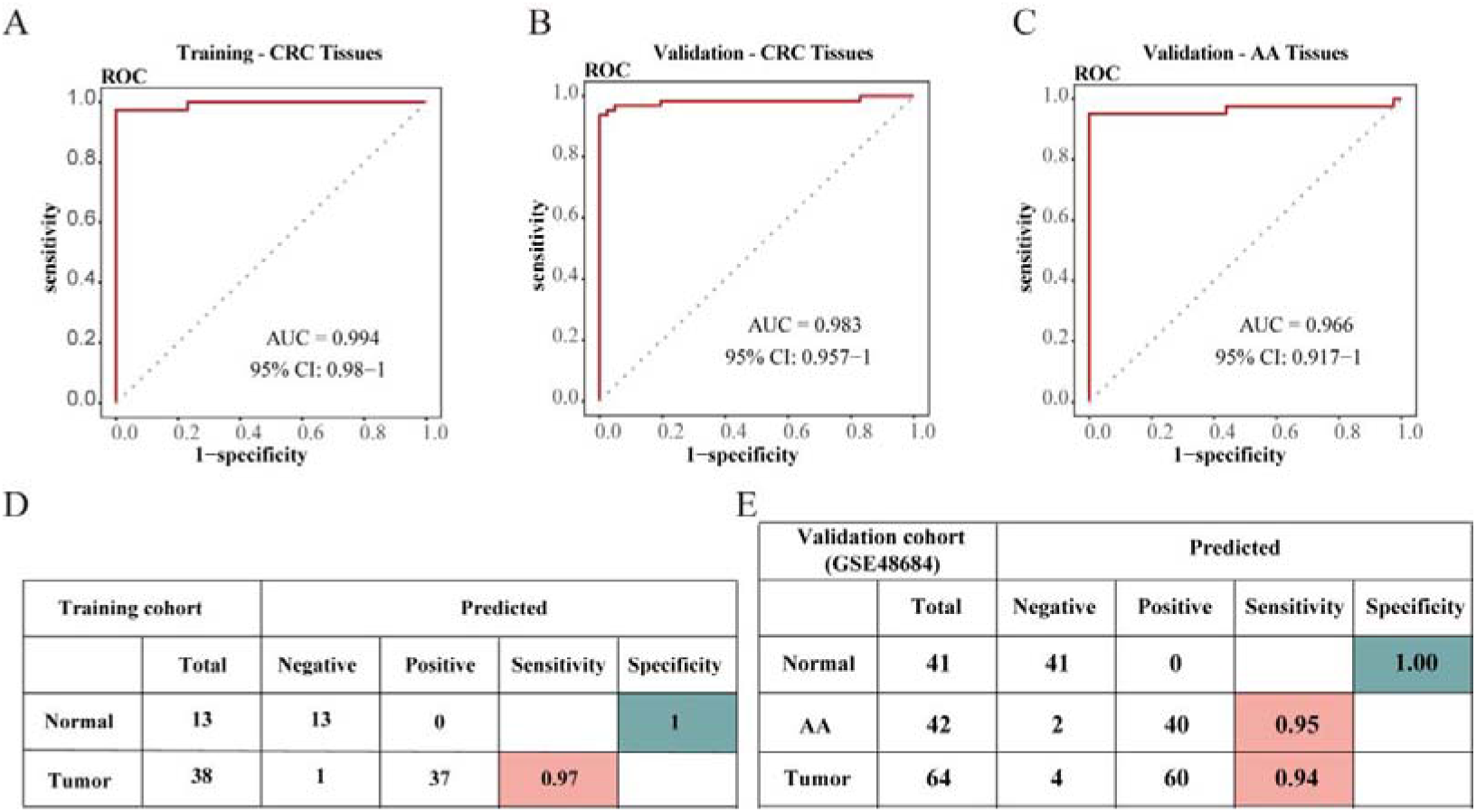
Development and validation of tissue diagnosis model based on the 27 CRC-specific DMRs. (A) The use of ROC curve analysis to assess the performance of the tissue diagnostic model in differentiating CRC patients from normal individuals of the training cohort (51 tissues we collected). (B-C) The use of ROC curve analysis to assess the performance of the tissue diagnostic model in differentiating CRC patients from normal individuals(B) and distinguish AA from Normal subjects (C) in tissue validation cohort 1(GSE 48684 dataset). (D-E) The sensitivity and specificity of the diagnosis model in the tissue training(D) and validation cohort 1(E).

**Figure 3.**
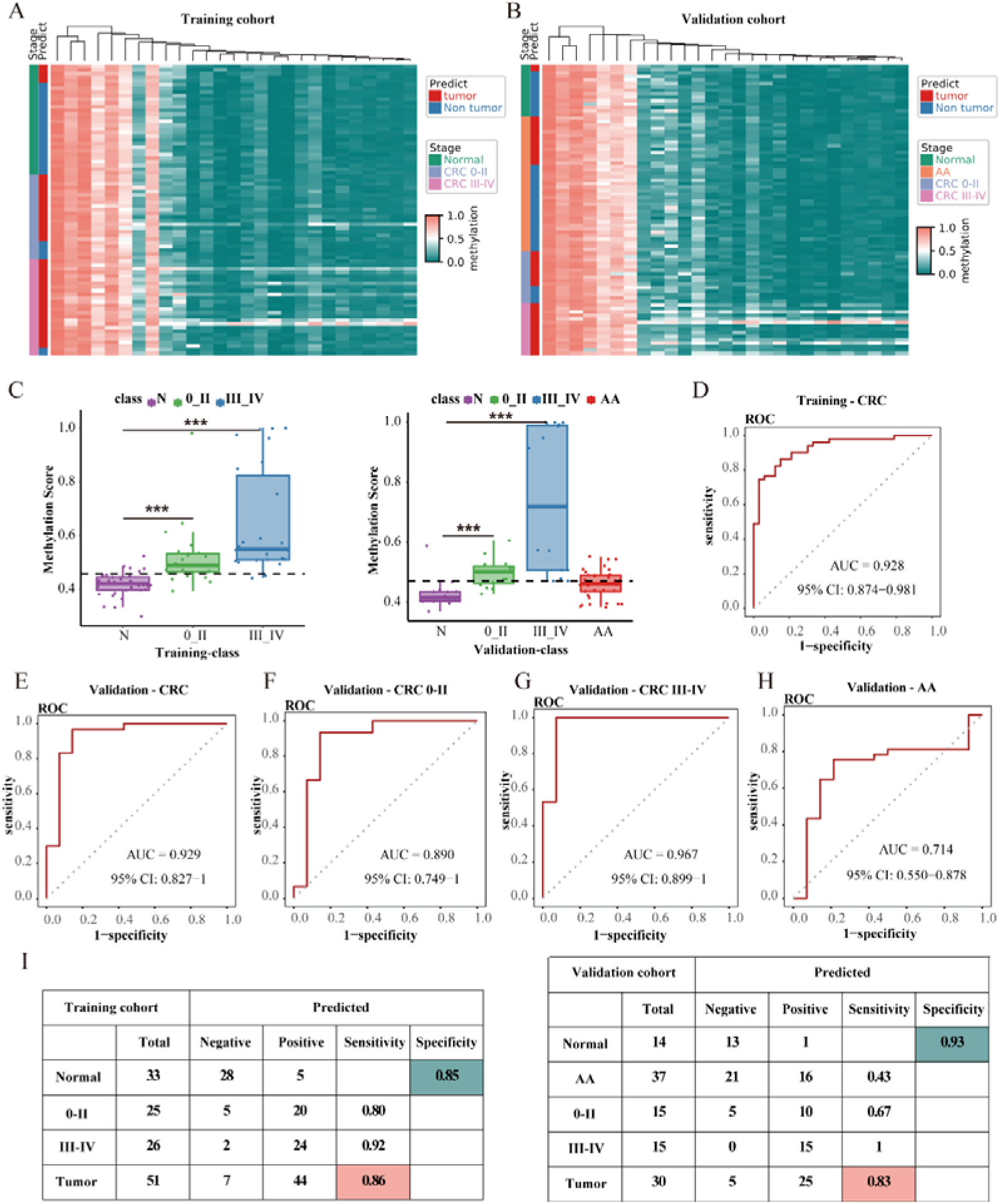
Development and validation of a plasma diagnosis model based on the 27 CRC-specific DMRs. (A-B) Heatmap illustrating the DMRs between CRC and advanced adenoma and healthy controls in the training (A) and validation cohort (B). (C) Methylation scores of Normal、AA and CRC individuals generated from the diagnosis model in the training and validation cohort (The dashed line represents the cutoff value). (D)The use of ROC curve analysis to assess the performance of the diagnosis model in differentiating CRC patients from normal individuals of the training cohort. (E) The use of ROC curve analysis to evaluate the performance of the diagnosis model in differentiating CRC patients from normal individuals of the validation cohort. (F-H) In the validation cohort, the performance of the diagnosis model in differentiating early-stage CRC (0-II) from normal individuals (F); in differentiating advanced-stage CRC (III-IV) from normal individuals (G); in differentiating AA from normal individuals (H). (I) The sensitivity and specificity of the diagnosis model stratified by stages in the training and validation cohort. Abbreviations: DMRs, differential methylated regions; ROC, receiver operating characteristic; AA, advanced adenoma; CRC, colorectal cancer.

(Figure 3E-H). The sensitivities of the CRC diagnosis model in the training cohort for stages 0-II, III-IV, and all CRC stages were 80%, 92%, and 86%, respectively, with a specificity of 85% (Figure 3I). The CRC diagnosis model demonstrated a specificity of 93% in the validation cohort, with sensitivities for AA, stages 0-II, III-IV, and all CRC stages of 43%, 67%, 100%, and 83%, respectively (Figure 3I). We further tested the early diagnostic performance of the model in an external validation cohort, and the results showed that the model achieved a sensitivity of 52% for AA and 48% for NAA (Figure 4A-E). Compared to the traditional tumor marker CEA, the model demonstrated higher sensitivity in detecting early-stage colorectal cancer and adenomas (Table s7). The plasma diagnosis model also serves to discern the specific staging of CRC patients. ROC curve analysis on the validation cohort revealed an AUC of 0.813 (0.656–0.970) for discriminating between CRC 0-II and CRC III-IV, and an AUC of 0.799 (0.693–0.905) for distinguishing between AA and CRC (Figure s5A-C). Kaplan-Meier survival curves revealed a significant reduction in overall survival (OS) for CRC III-IV patients compared to CRC 0-II patients identified by this diagnosis model (Figure s5D-E).

**Figure 4.**
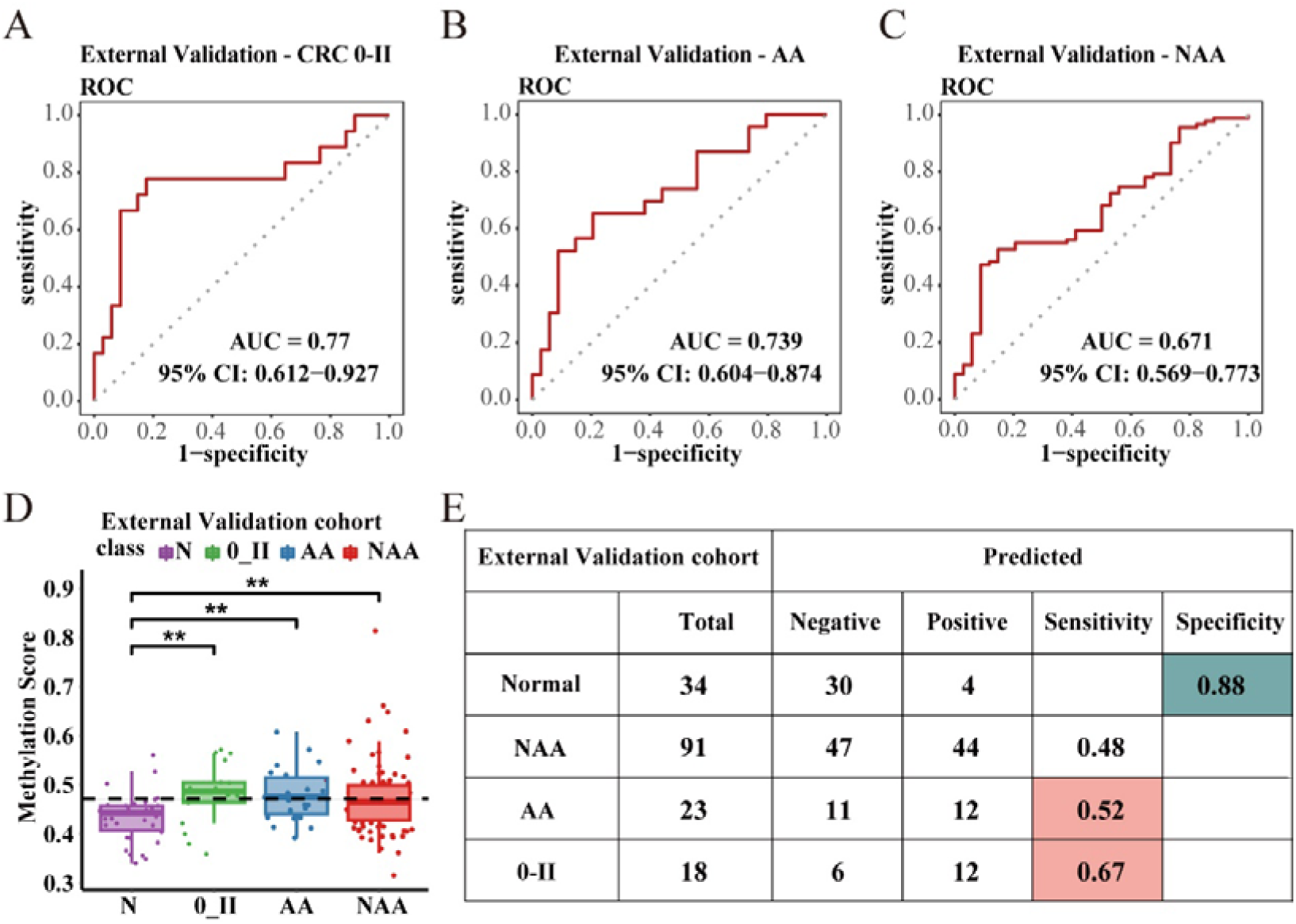
External validation of plasma diagnosis model. (A-C) In the external validation cohort, the performance of the diagnosis model in differentiating early-stage CRC (0-II) from normal individuals (A); in differentiating AA from normal individuals (B); in differentiating NAA from normal individuals (C). (D) Methylation scores of Normal、NAA、AA and CRC (0-II) individuals generated from the diagnosis model in the external validation cohort (The dashed line represents the cutoff value). (E) The sensitivity and specificity of the diagnostic model for NAA, AA, and CRC in the external validation cohort.

### Development and validation of CRC metastasis and prognosis models

To investigate the clinical relevance of our methylation signature, we first correlated the plasma methylation scores with clinicopathological parameters. The scores were significantly associated with tumor stage and metastasis (*P* < 0.001, Figure 5A-B) but not with age, sex, or lesion location (*P* > 0.05, Figure s6A-C). Based on these associations, we developed models to predict metastasis and prognosis using the 27-DMR panel. The metastasis model demonstrated excellent performance, achieving an AUC of 0.969 (95% CI: 0.926-1) in the training cohort and 0.955 (95% CI: 0.878-1) in the validation cohort (Figure 5C). Correspondingly, patients with metastatic disease (M1) had significantly higher methylation scores than those without (M0) and exhibited shorter overall survival (OS) (Figure s6D-F). The prognosis model also showed high accuracy, with AUCs of 0.883 (95% CI: 0.778-0.988) in the training set and 0.867 (95% CI = 0.728-1) in the validation set (Figure 5D). Using an optimal cutoff, the prognosis model effectively stratified patients into high-and low-risk groups. Kaplan-Meier analysis revealed that patients in the high-risk group had significantly worse OS (Figure 5E). Critically, this high-risk group encompassed nearly all patients who developed distant metastases or died during follow-up (Figure 5F-I). To confirm the score’s independent prognostic value, we performed a multivariate Cox regression analysis, which showed that the 27-DMR methylation score was an independent predictor of OS (Table S6). These findings establish that our models can successfully stratify patients, identifying those who may require more aggressive treatment.

**Figure 5.**
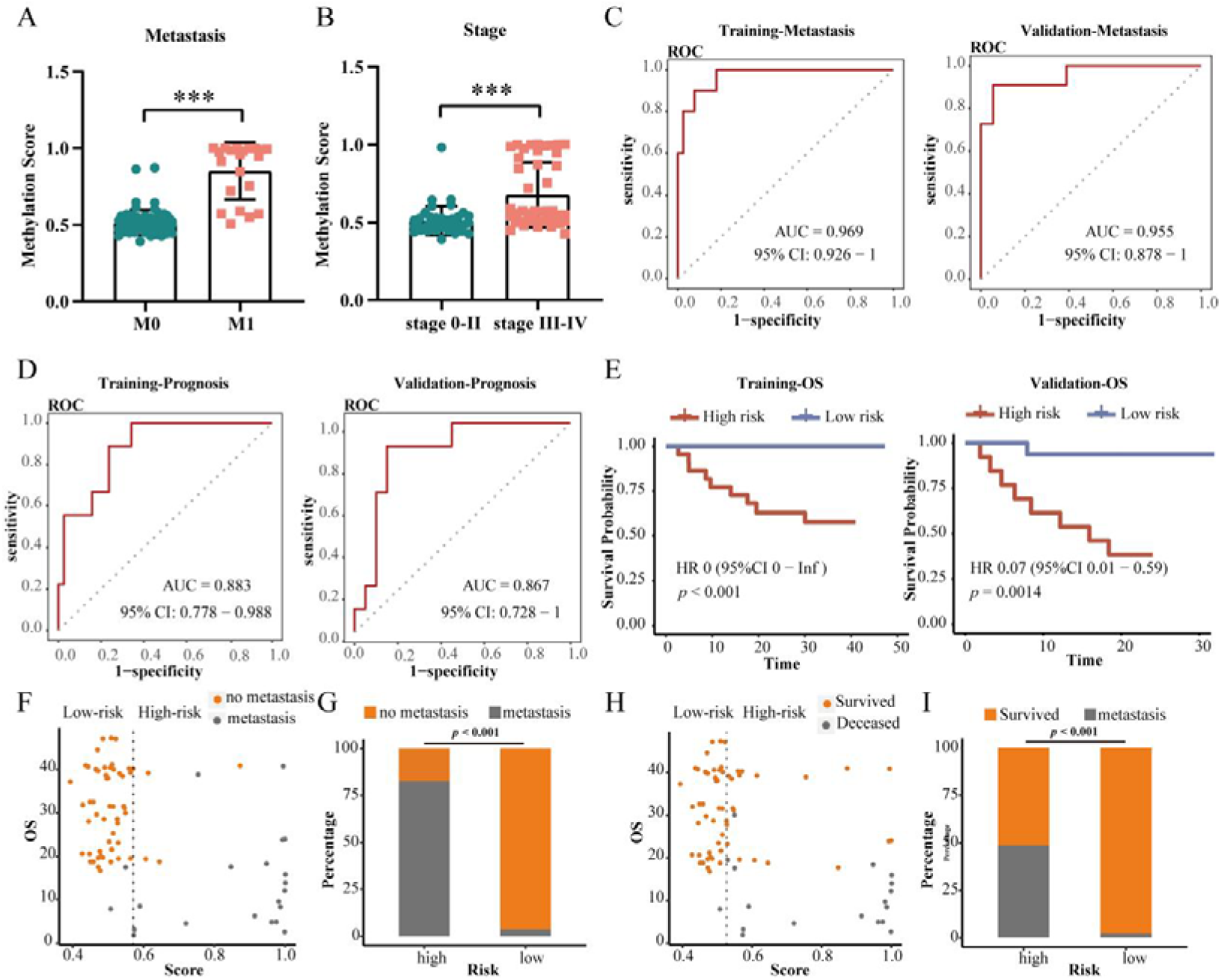
Development and validation of a plasma-based metastasis model and a plasma-based prognosis model. (A-B) The methylation scores generated by the diagnosis model for CRC patients across various M stages (A) and different disease stages (B). (C) ROC curve analysis was performed to assess the performance of 27 DMRs methylation scores generated by the diagnosis model in distinguishing M1-CRC patients from M0-CRC patients in the training (AUC = 0.969) and validation (AUC = 0.955) cohorts. (D) ROC analysis evaluates the performance of 27 DMRs methylation scores generated by the diagnosis model in predicting the prognosis of CRC patients. (E) Kaplan-Meier survival curves comparing OS between the high-risk CRC group and low-risk CRC group of prognosis model in the training cohort and validation cohort. (F, H) Metastasis and prognosis prediction of the training and validation patitients (n = 88). The black dotted line at the cutoff value divides the patients into high-risk and low-risk groups. Yellow circles and gray circles represent patients without distant metastasis and with distant metastasis, respectively (F). Yellow circles and gray circles represent patients survived and deceased, respectively (H). (G, I) The proportion of metastasis and deceased is higher in the high-risk group. The p-value was calculated using a two-sided Fisher’s exact test. Abbreviations: DMRs, differential methylated regions; ROC, receiver operating characteristic; M1-CRC, colorectal cancer with distant metastasis; M0-CRC, colorectal cancer without distant metastasis; AUC, the area under the curve.

### Distinct Release Dynamics of Methylation Markers from Tissue to Plasma

To understand the biological basis for the model’s performance characteristics, we compared methylation scores between paired tissue and plasma samples across the spectrum of disease progression. We found that in tissue, the 27-DMR signature was already strongly present at the precancerous adenoma (AA) stage (Figure 6A). However, in plasma, the signal intensity increased in a stepwise manner, becoming most prominent in advanced-stage CRC (Figure 6B). This discrepancy suggests that the release of cfDNA markers into circulation is stage-dependent. Indeed, a subset of DMRs was detectable in plasma even in early-stage disease, and these markers were associated with genes involved in cellular secretion pathways (Figure 6C, s6G). In contrast, other DMRs were only detected in the plasma of patients with advanced CRC. This suggests a dual mechanism of release: limited, active secretion of certain markers from early lesions, followed by massive, passive release from widespread cell death in larger, necrotic tumors. This dynamic explains why the diagnostic sensitivity of the plasma test is highest for advanced-stage disease.

**Figure 6.**
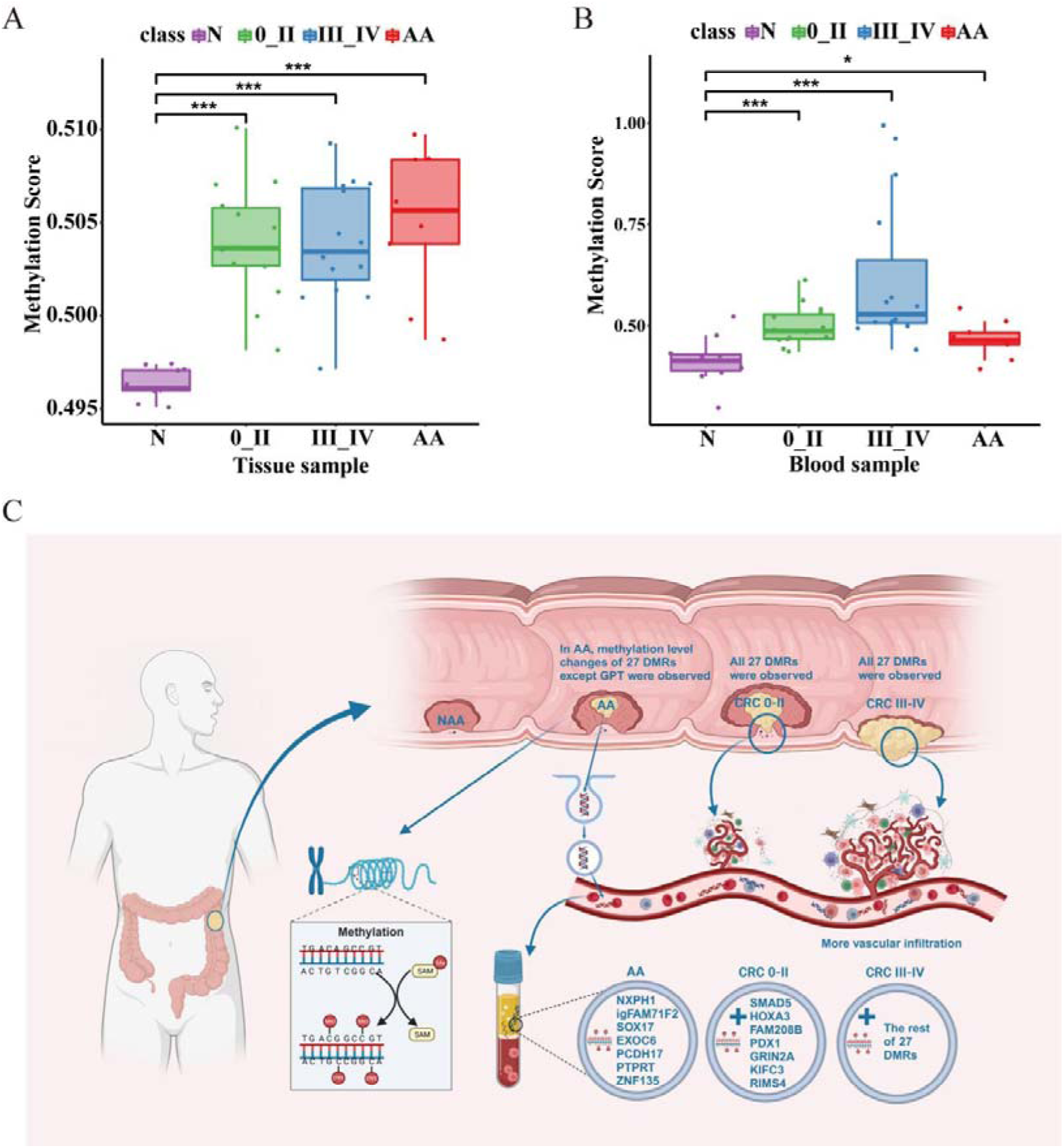
Methylation scores of paired tissue and plasma samples. (A) The methylation scores of tissues from Normal, AA, CRC 0-II, CRC III-IV individuals. (B) The methylation scores of blood samples from Normal, AA, CRC 0-II, CRC III-IV individuals. (C) The mechanism diagram interpreting the methylation scores changes of tissue and plasma across different populations.

## Discussion

Liquid biopsy of cfDNA has become an ideal clinical detection method because it is minimally invasive and easily sampled. However, a key challenge in liquid biopsy is confirming that cfDNA markers truly originate from tumor tissue. We addressed this by implementing a rigorous, multi-stage screening strategy that began with TCGA data, was validated in our own CRC tissue samples, and was finally optimized in plasma. This ensures that our 27-DMR signature is not only statistically powerful but also biologically rooted in CRC pathobiology. In this study, we implemented a rigorous, multi-stage screening strategy that began with integrating TCGA tissue data with self-collected tissue methylation data to identify CRC-related methylation sites, was validated in our own CRC tissue samples, and was finally optimized in plasma. This ensures that our 27-DMR signature is not only statistically powerful but also biologically rooted in CRC pathobiology. Our resulting diagnostic model, built upon these tissue-of-origin-confirmed markers, demonstrates significant potential for early diagnosis, staging, metastasis and prognostic assessment.

A critical benchmark for any new CRC screening tool is its performance against existing methods, especially in detecting precancerous lesions. For instance, the FDA-approved circulating methylated SEPT9 DNA (mSEPT9) offers a specificity of 91.5% but provides limited sensitivities of 11.2%, 35.0%, 63.0%, 46.0%, and 77.4% for AA and CRC stages I-IV, respectively (^16^). While more recent cfDNA methylation models have improved overall CRC detection, their ability to detect precancerous lesions can still be modest; one such study reported a sensitivity of 33.3% for adenomas (^17^). In this context, our compact 27-marker plasma model demonstrates a highly competitive performance, achieving an overall sensitivity of 83.3% and a specificity of 92.9% for CRC. Most notably, our model shows a substantial improvement in detecting high-risk precancerous lesions, with sensitivities for advanced adenomas (AA) and non-advanced adenomas (NAA) reaching 52.2% and 48.4%, respectively. This represents a nearly five-fold increase in sensitivity for advanced adenomas compared to the mSEPT9 test (^16, 17^). This enhanced capability positions our model as a promising new tool for CRC screening in high-risk populations, enabling earlier intervention before the onset of malignancy.

In plasma, we observed the methylation scores of 27 DMRs gradually increase with CRC progression. It is possibly due to less vascular infiltration in early CRC (^18, 19^). On the other hand, the ctDNA detected in the blood of early-stage CRC is largely derived from tumor cells actively secreting into the bloodstream, making it challenging to be precisely captured with current detection technologies due to the limited quantity. Conversely, ctDNA in the blood of late-stage CRC primarily emanated from apoptotic and necrotic tumor cells, resulting in a larger quantity that facilitates easier detection. However, in reality, the methylation scores of the 27 DMRs have indeed exhibited noticeable changes in AA and early cancerous tissues. Additionally, in the tissue diagnosis model using the 27 DMRs, the sensitivity was 94% and the specificity was 100%. Therefore, we have grounds to believe that the 27 DMRs detected in plasma originate from CRC tumor tissues. With the implementation of more sensitive detection methods, the performance of these 27 DMRs in early CRC plasma diagnosis is likely to be further enhanced.

Beyond early detection, cfDNA analysis is a valuable tool for risk stratification and monitoring for recurrence in CRC (^20^). However, many current approaches require serial postoperative testing to track disease progression (^15, 21^). A key advantage of our model is its ability to provide significant prognostic information from a single, preoperative blood sample. Our findings reveal that the 27-DMR methylation score progressively increases with CRC stage advancement, a feature with direct clinical utility (^22, 23^). Firstly, this allows for effective preoperative staging, as the model can distinguish early-stage (0–II) from late-stage (III–IV) CRC with high accuracy (AUC = 0.813), offering a simple, non-invasive method to help guide initial treatment planning. Secondly, and perhaps more significantly, the preoperative score is a powerful predictor of patient prognosis. It was strongly associated with distant metastasis (AUC = 0.955), enabling the stratification of patients into high-and low-risk groups before surgery even begins. This capability is particularly valuable as it provides crucial prognostic insights that can inform perioperative management—such as identifying patients who may require more aggressive treatment or intensive surveillance—from a single baseline measurement, potentially reducing the reliance on continuous postoperative monitoring.

The treatment strategies for CRC differ significantly across various stages. Molecular stratification approaches for CRC patients are on the rise, and concurrently, the clinical application of biomarkers to determine treatment decisions is gradually gaining traction (^24^). Studies have demonstrated the precise identification of T1 CRC patients at risk of lymph node metastasis using a specific set of miRNAs, thereby potentially mitigating unnecessary overtreatment (^25, 26^). Our CRC diagnosis model, with an AUC of 0.813 for distinguishing CRC 0-II from CRC III-IV, could also serve as a potent, straightforward, and cost-effective preoperative screening/detection method, guiding patients to select more appropriate treatment plans.

DNA methylation regulates gene transcription, guiding the progression from normal mucosa to AA and ultimately CRC. This process involves silencing tumor suppressor genes and activating oncogene transcription (^27^). The clinical performance of our panel is underpinned by the biological functions of its associated genes, many of which are known to be involved in CRC pathogenesis. For instance, high methylation and diminished expression of transmembrane protein 240 (TMEM240) regulate CRC cell proliferation, predicting poor prognosis (^28^). The transcription factor homeobox A3 (HOXA3) activates aerobic glycolysis, promoting tumor growth (^29^). KIFC3 controls mitotic spindle assembly initiation (^30^). Moreover, several genes are implicated in tumor metastasis, such as NOVA alternative splicing regulator 1 (NOVA1), which promotes CRC migration by activating the Notch pathway (^31^). Protein tyrosine phosphatase receptor type T (PTPRT) contributes to early CRC dissemination (^32^). SPARC-related modular calcium binding 2 (SMOC2) serves as the distinctive signature of cancer stem cells (CSCs) in CRC and promotes epithelial-to-mesenchymal transition (EMT) (^33, 34^). Increased methylation and expression of pancreatic and duodenal homeobox 1 (PDX1) facilitate CRC invasion and migration(^35^). The correlation we observed between the methylation levels of our 27 DMRs and the transcription levels of their respective genes in TCGA datasets further supports their functional relevance. However, the mechanisms by which alterations in specific DNA methylation impact gene expression are intricate. Certain transcription factors selectively recognize sequences with methylated CpG (mCpG) and influence the expression of multiple genes(^36^). Further exploration of the potential functional mechanisms of these DNA methylation markers may deepen our understanding of the molecular processes underlying CRC development, offering promising therapeutic targets. Despite the promising results, we acknowledge several limitations in this study. Firstly, although obtained from multiple institutions, the overall sample size is relatively small. Therefore, the cfDNA methylation model should be further validated in larger, prospective clinical trials including a larger and more balanced set of precancerous lesions. Secondly, a prospective study is necessary to directly compare or combine the cfDNA methylation model with clinically established protein markers such as CEA and CA19-9, and Fecal Immunochemical Test (FIT) to determine its ultimate clinical utility.

In conclusion, our study introduces a novel, tissue-validated 27-DMR cfDNA methylation signature that effectively serves three critical clinical needs: early detection of CRC, including high-risk precancerous lesions; preoperative staging; and prognostic risk stratification for metastasis. As a robust, convenient, and cost-effective detection method, the preoperative application of this biomarker panel has the potential to facilitate more informed clinical decision-making and significantly improve the management of patients with colorectal cancer.

## AUTHOR CONTRIBUTIONS

Yuqi He and Jianqiu Sheng conceived and designed the study. Lang Yang, Fangli Men, Jianwei Yu, Xianzong Ma, Junfeng Xu, Yangjie Li, Ju Tian, Hui Xie, Qian Kang, Linghui Duan, Xiang Yi, Wei Guo, Ni Guo collected the samples and curated the date. Xueqing Gong, Lingqin Zhu, Lang Yang, Fangli Men, Jianwei Yu, Shuyang Sun, Chenguang Li, Xianzong Ma, Junfeng Xu, Yangjie Li, Xiang Yi, Wei Guo, Ni Guo, Youyong Lu, Joseph Leung, Yuqi He and Jianqiu Sheng analysed the data. Lingqin Zhu wrote the manuscript with the assistance of Youyong Lu, Joseph Leung, Yuqi He and Jianqiu Sheng.

## Data Availability

All data produced in the present study are available upon reasonable request to the authors.

https://www.ncbi.nlm.nih.gov/geo/query/acc.cgi?acc=GSE48684

https://portal.gdc.cancer.gov/projects/TCGA-READ

## ACKNOWLEDGEMENTS

This research was funded by grants from the National Natural Science Foundation of China (Grant no. 82273245), the Capital’s Funds for Health Improvement and Research (Grant no. 2022-1-5082), and the Beijing Natural Science Foundation (Grant no. 7212107), Sponsored by Dongying City Natural Science Foundation (Grant no. 2023ZR026), Sponsored by Longyan City Science and Technology Plan Project (Grant no. 2022LYF17082).

## DATA AVAILABILITY STATEMENT

The data has been deposited in the National Genomics Data Center (NGDC) and is accessible under the accession number OMIX009128. All of the data supporting this work will be made available from the corresponding author upon reasonable request.

## ETHICS APPROVAL AND CONSENT TO PARTICIPATE

The study was conducted in accordance with the Declaration of Helsinki, and approved by the Ethics Committee of the Seventh Medical Center of PLA General Hospital (Approval No. 2016-70, 2020-78), Dongying People’s Hospital (Approval No. DYYW-2019-002-01), and the First Hospital of Longyan, Fujian Medical University (Approval No. 2021-k0001). Informed consents were obtained from all participants involved in the study.

## Online Supplementary Material

**Contents**

## Supplementary Methods

**Supplementary Figure s1-s6**

Supplementary Figure 1(Figure s1). Screening for CRC-specific DMCs based on TCGA data.

Supplementary Figure 2(Figure s2). DMCs selection in tissues.

Supplementary Figure 3(Figure s3). Selection of methylation markers.

Supplementary Figure 4(Figure s4). The relationship between the methylation levels of the 27 DMRs and the transcription levels of their respective genes in TCGA datasets.

Supplementary Figure 5(Figure s5). Development and validation of a plasma diagnosis model for discerning the specific staging of CRC patients.

Supplementary Figure 6(Figure s6). The construction of the metastasis model.

**Supplementary Table s1-s6**

Table s1 Explanation of specific phrase and abbreviation Table s2 Patient characteristics of the 15 paired tissues

Table s3 Patient characteristics of the 64 participants (Tissues) Table s4 Patient characteristics of the blood cohort

Table s5. Annotation information for 27 regions

Table s6 Univariate and multivariate analysis of methylation scores and clinical parameters.

Table s7 Comparison of diagnostic performance between the methylation score and CEA for early-stage colorectal cancer and adenomas in the external validation cohort.

## Supplementary Methods

### The inclusion and exclusion criteria

Inclusion criteria included individuals aged over 18, with healthy control subjects having negative results from colonoscopy or pathology, and no history of other cancers. Patients with colorectal cancer and precancerous lesion were required to have a confirmed diagnosis with colonoscopy and/or pathology, and without concurrent malignancies involving other systems or organs. Blood was collected from participants before any form of treatment (surgery, radiotherapy, or chemotherapy).

### CRC-specific methylation markers selection

Using the following methodology, tissue-specific methylation markers were identified for CRC. Initially, ChAMP differential analysis software was used to analyze CRC methylation data (450k chip methylation data) from TCGA, filtering out differentially methylated sites between CRC and control (Δβ>0.50, FDR<0.05). The top 500 sites, contingent on the extent of differential methylation, were earmarked as candidate marker sites. Merging TCGA CRC methylation data with transcriptome data, 477 hypermethylated sites (Δβ>0.20, FDR<0.05) in the TSS and First Exon regions of downregulated genes (log2FC<−1 and FDR<0.05) were selected. DNA methylation data encompassing CRC, esophageal cancer, gastric cancer, lung cancer, liver cancer, and breast cancer were retrieved from the TCGA database. Using the ChAMP differential analysis software, 499 CRC-specific methylation sites were identified (CRC, Δβ>0.20, FDR<0.05; for other cancers, Δβ≤0.20). The amalgamation of the outcomes from the aforementioned three stages, supplemented by the inclusion of previously reported CRC-related methylation sites from the literature, yielded a list of 1438 potential differential methylated CpG sites (DMCs). Subsequently, DNA methylation of 15 pairs of tissue samples from Chinese CRC patients (comprising 5 pairs of advanced adenoma tissues and 10 pairs of CRC tissues) were sequenced by Roche Nimble Gen’s SeqCap Epi method, which encompasses approximately 80Mb of Roche methylation probes, covering 5.5 million methylation sites, including all sites from the Illumina 450K chip. The top 1400 DMCs were selected based on adjusted *P* value (Δβ > 0.20, FDR < 0.05). These 1400 DMCs were then amalgamated with the previously identified 1438 sites, and custom-designed probes were synthesized. By testing the 89 samples (13 normal tissues and 38 paired CRC tissues) with the special probes, the DMCs between tumors and adjacent tissues, as well as between cancer and normal tissues were identified (Δβ > 0.20, FDR < 0.05). Through the intersection of these identified sites and the amalgamation of those in close proximity, 404 CRC-specific DMRs were ultimately identified.

Subsequently, the methylation levels of 77 plasma samples (13 Normal, 15 NAA, 12 AA, 37 CRC) were assessed using the Wilcox Differential Analysis (Δβ > 0.01, FDR < 0.05). To further reduce methylation markers, 500 random iterations were performed, extracting 75% of the samples from the training set each time. Subsequently, the Least Absolute Shrinkage and Selection Operator (LASSO) regression analysis was employed to filter markers. Ultimately, 27 regions that appeared in more than 50 iterations were chosen as plasma diagnostic markers. The relationship between the methylation levels of the 27 DMRs and the transcription levels of their respective genes was compared by Spearman correlation coefficients.

### DNA extraction from tissues and plasma

DNA extraction from AA, CRC, and healthy colon tissues was carried out using the TIANamp Genomic DNA Kit (DP304, TIANGEN, Beijing, China). Before DNA extraction, tissue pathology evaluation confirmed the presence of tumor cells in CRC tissue samples. We collected 10 milliliters of whole blood from each participant. First, they were centrifuged at 1400×g for 25 minutes at 4LJ temperature. Then, the upper layer was transferred and centrifuged again at 14000 ×g for 10 minutes at 4LJ temperature to obtain plasma. The MagMAX™ Cell-Free DNA Isolation Kit (A29319, Thermo Fisher, Waltham, MA, USA) was utilized for cfDNA extraction from plasma, with special attention to avoiding repeated freeze-thaw cycles to prevent cfDNA degradation. The concentration of the isolated cfDNA was analyzed by fluorometric quantification using the Qubit Fluorometer 4.0 (Thermo Fisher, Waltham, MA, USA) and the Qubit dsDNA HS Assay Kit (Thermo Fisher). The fragment size profiles of cfDNA were assessed using the 2100 bioanalyzer (Agilent Inc., Santa Clara, CA, USA) with the Agilent High-Sensitivity DNA Kit. CfDNA with a yield < 5 ng or exhibiting quality issues was excluded.

### Library construction

Up to 1000 ng of gDNA was fragmented with Covaris M220 and used to construct an indexed library using KAPA HyperPrep Kit, PCR free. Then follow the steps of the EZ DNA Methylation Lightning Kit (Zymo Research; Irvine, CA) for the bisulfite conversion of the DNA library. Up to 5 ng of plasma cfDNA was subjected to bisulfite conversion using the EZ DNA Methylation Lightning Kit (Zymo Research; Irvine, CA), converted cfDNA was used to prepare indexed sequencing library using Accel-NGS Methyl-Seq DNA library preparation kits (Swift BioSciences; Ann Arbor, MI).

### Targeted bisulfite sequencing using the SeqCap Epi Enrichment System

Libraries were quantified using the Qubit dsDNA HS Assay Kit before pooling 3 (cfDNA) or 4 (gDNA) individual libraries per capture of SeqCap Epi CpGiant probes from Roche Nimblegen. Hybridization and capture were performed using the HyperCap Target Enrichment Kit from Roche. Pooled libraries were sequenced on an Illumina Hiseq X Ten using paired-end 150-bp reads.

### Targeted bisulfite sequencing using the Twist Target Enrichment System

Libraries were quantified using the Qubit dsDNA HS Assay Kit before pooling 4 or 8 individual libraries per capture of Twist Human Methylome Panel or Twist Methyl Custom Panel respectively. Hybridization and capture were performed using the Fast Hybridization Target Enrichment kit from Twist Bioscience. Pooled libraries were sequenced on an Illumina Novaseq 6000 or Hiseq X Ten using paired-end 150-bp reads.

### Methylation data processing

The raw image data files acquired through high-throughput sequencing were subjected to base calling using the bcl2fastq software, resulting in the conversion to raw sequencing data stored in the fastq file format. Rigorous quality control measures were applied to the on-machine raw data, facilitating the exclusion of reads of inferior quality. Bismark (Krueger, 2011, invoking Bowtie2 at the core) was used for the alignment analysis of methylation data with the reference genome. After the acquisition of alignment results, Bismark (Krueger, 2011) was once again utilized for the identification of methylation sites. Differential methylation analysis for paired samples was conducted employing the paired t-test method, while unpaired samples underwent differential methylation analysis utilizing the wilcoxon rank sum method.

## Supplementary Figures

**Supplementary Figure 1.**
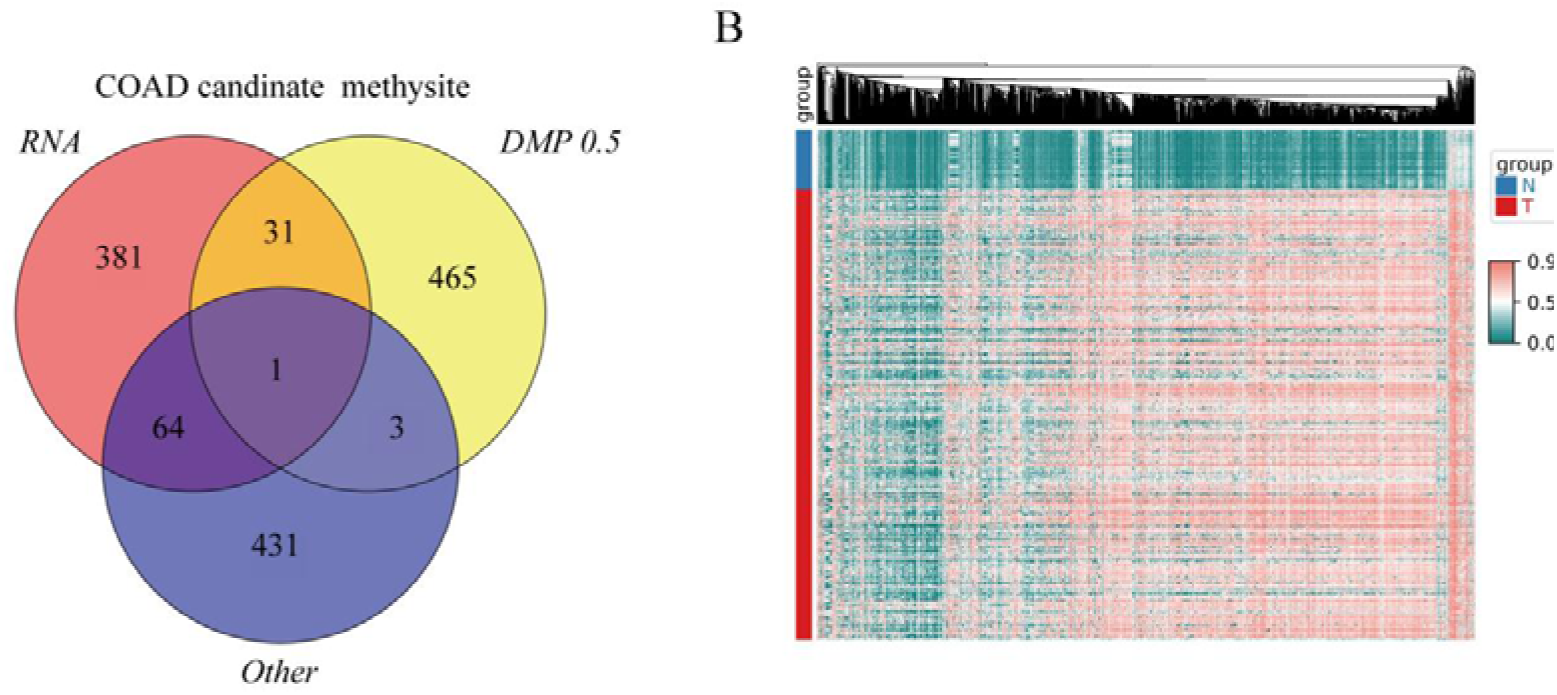
Screening for CRC-specific DMCs based on TCGA data. (A) Venn diagram of the results from three DMCs filtering methods based on TCGA datasets. (B) Heatmap illustrating the methylation level of the 1438 methylated sites by unsupervised clustering. Abbreviations: DMCs, differential methylated CpG sites; CRC, colorectal cancer.

**Supplementary Figure 2.**
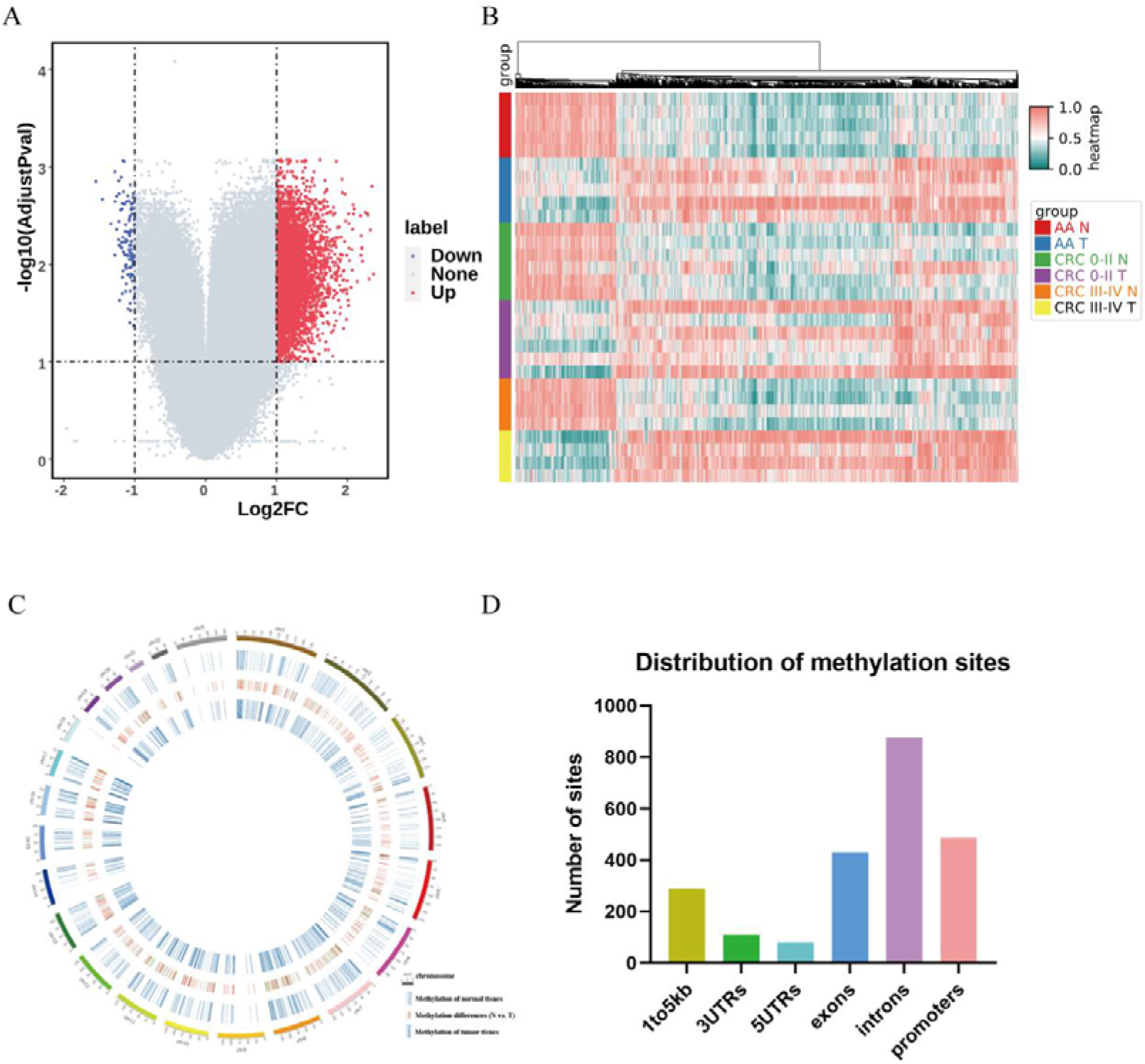
DMCs selection in tissues we collected. (A) Volcano plot illustrates the CRC specific DMCs. (B) Heatmap analysis of the top 1400 DMCs between CRC tissues and adjacent tissues by unsupervised clustering. (C) Circular plot of 1400 DMCs depicting methylation level differences between CRC and adjacent tissues. (D) Distribution of 1400 DMCs in gene functional regions. Abbreviations: DMCs, differential methylated CpG sites; CRC, colorectal cancer.

**Supplementary Figure 3.**
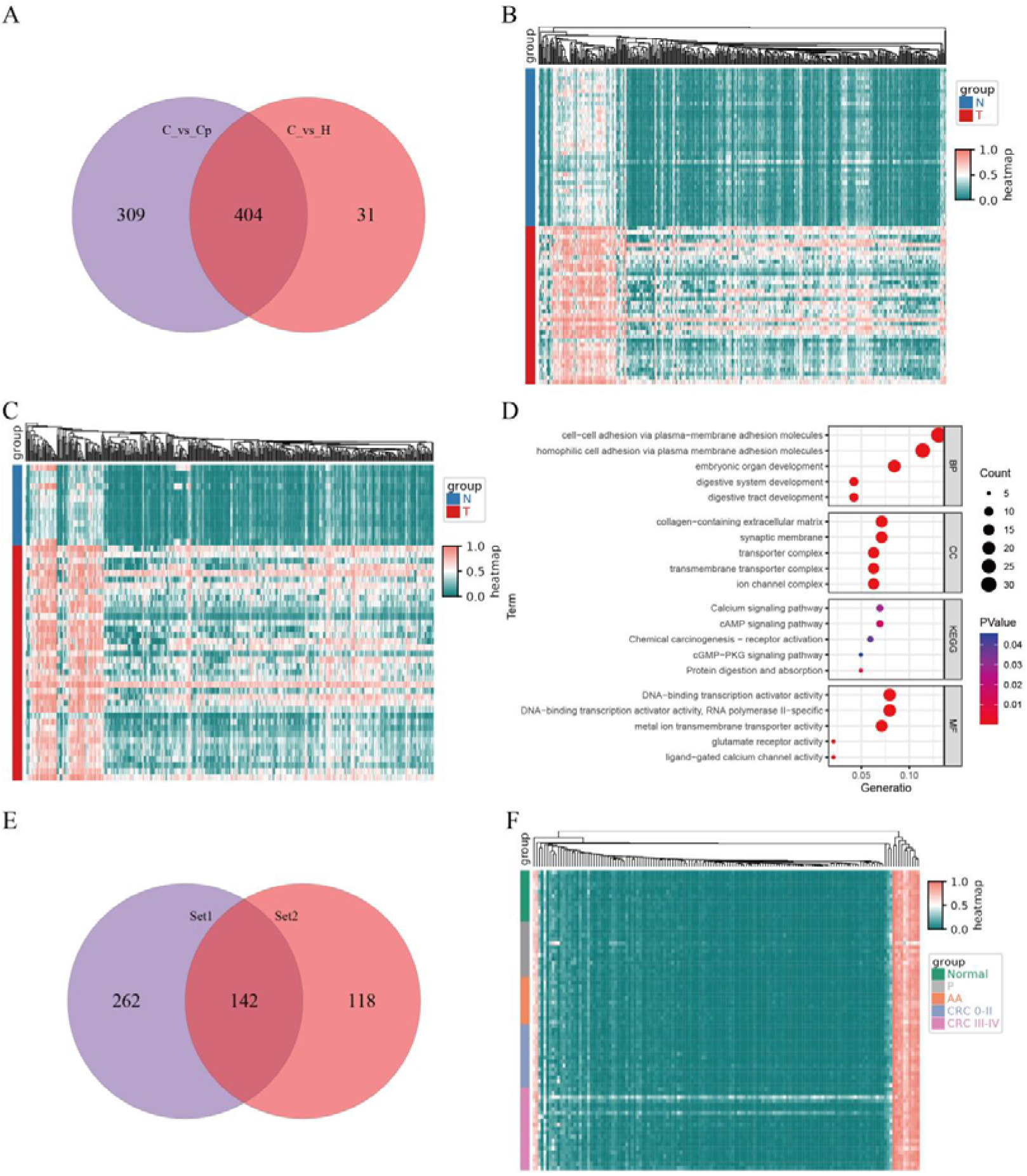
Selection of methylation markers. (A) The overlap of DMRs between CRC and adjacent tissues, as well as between CRC and normal tissues. (B) Heatmap illustrating the methylation level of the 404 DMRs in CRC and adjacent tissues. (C) The methylation level of the 404 DMRs in CRC and normal tissues. (D) The gene enrichment analysis for the 404 regions suggests their involvement in processes like cell-cell adhesion, digestive system development, and signaling pathways such as cAMP and cGMP. (E) Based on the results of plasma methylation detection, 142 DMRs were subsequently chosen for further analysis. (F) Heatmap illustrating the methylation levels of the 142 DMRs in the blood samples from normal individuals, those with AA, and patients with CRC. Abbreviations: DMRs, differential methylated regions; AA, adenomatous polyps; CRC, colorectal cancer.

**Supplementary Figure 4.**
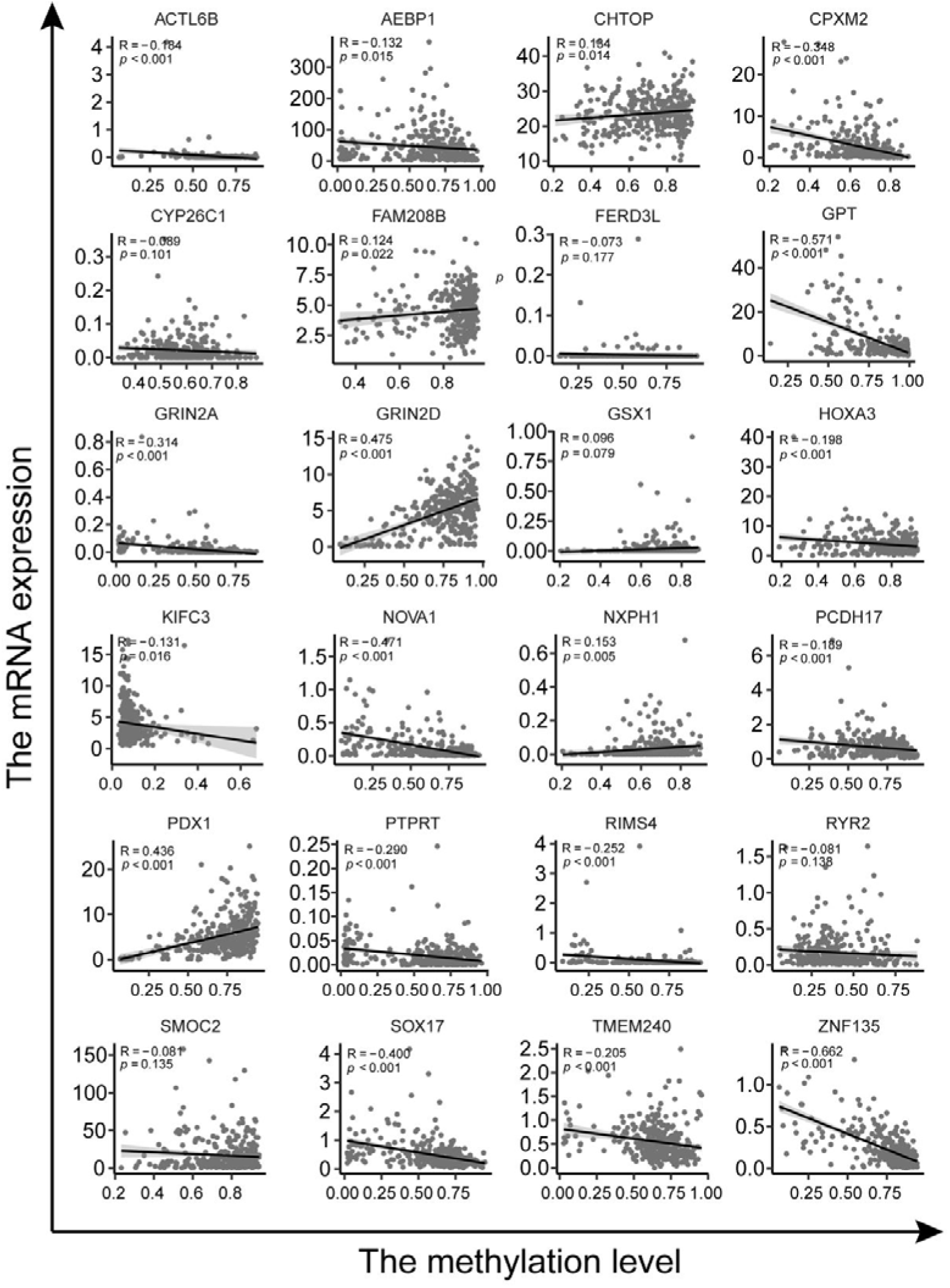
The relationship between the methylation levels of the 27 DMRs and the transcription levels of their respective genes in TCGA datasets.

**Supplementary Figure 5.**
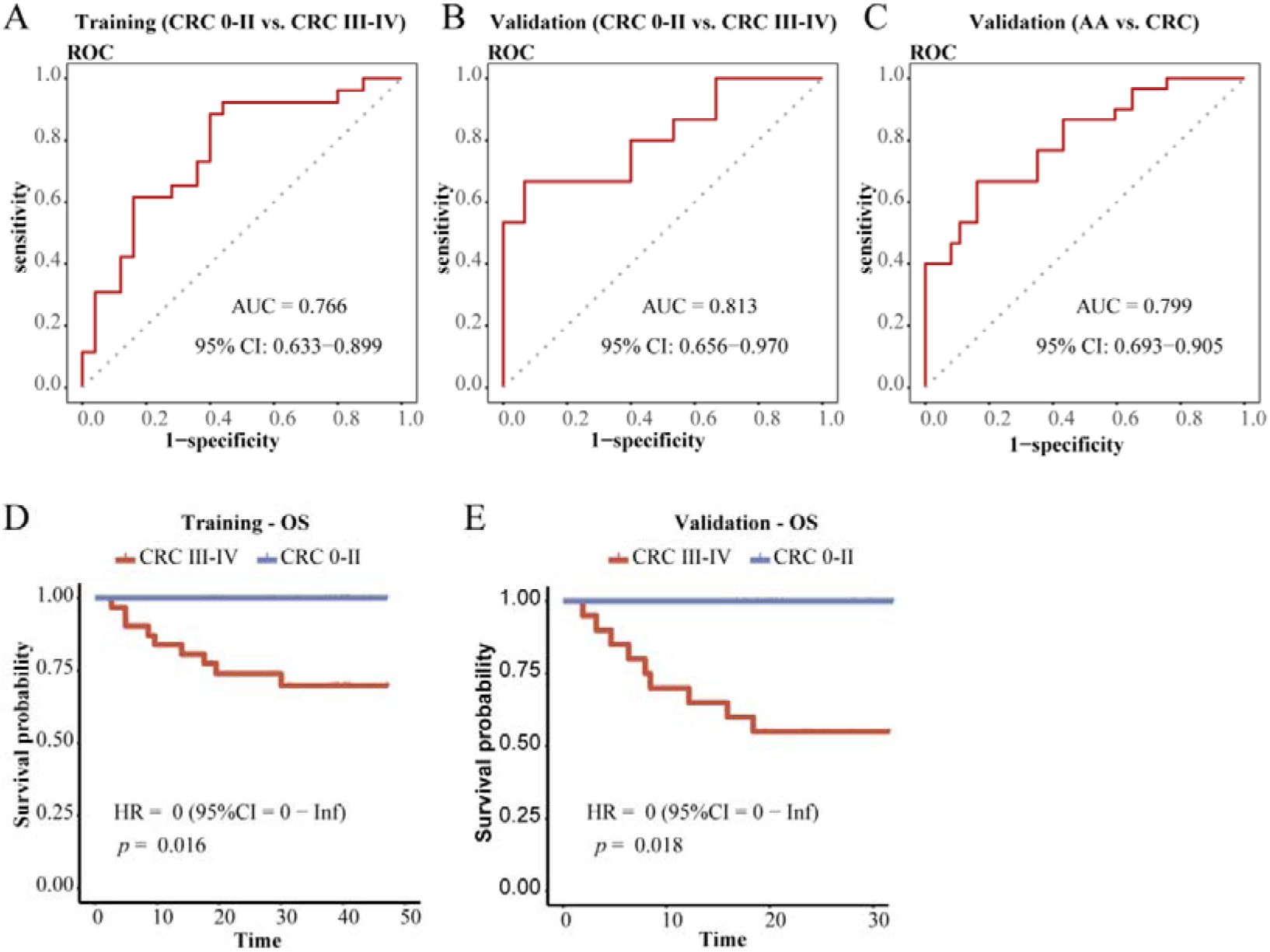
Development and validation of a plasma diagnosis model for discerning the specific staging of CRC patients. (A) The use of ROC curve analysis to assess the performance of the diagnostic model in differentiating CRC 0-II patients from CRC III-IV patients of the training cohort. (B-C) The use of ROC curve analysis to evaluate the performance of the diagnostic model in distinguishing CRC 0-II patients from CRC III-IV patients (B), AA from CRC patients (C) of the validation cohort. (D-E) Kaplan-Meier survival curves comparing OS between the CRC III-IV group and CRC 0-II group in the training cohort (D) and validation cohort (E).

**Supplementary Figure 6.**
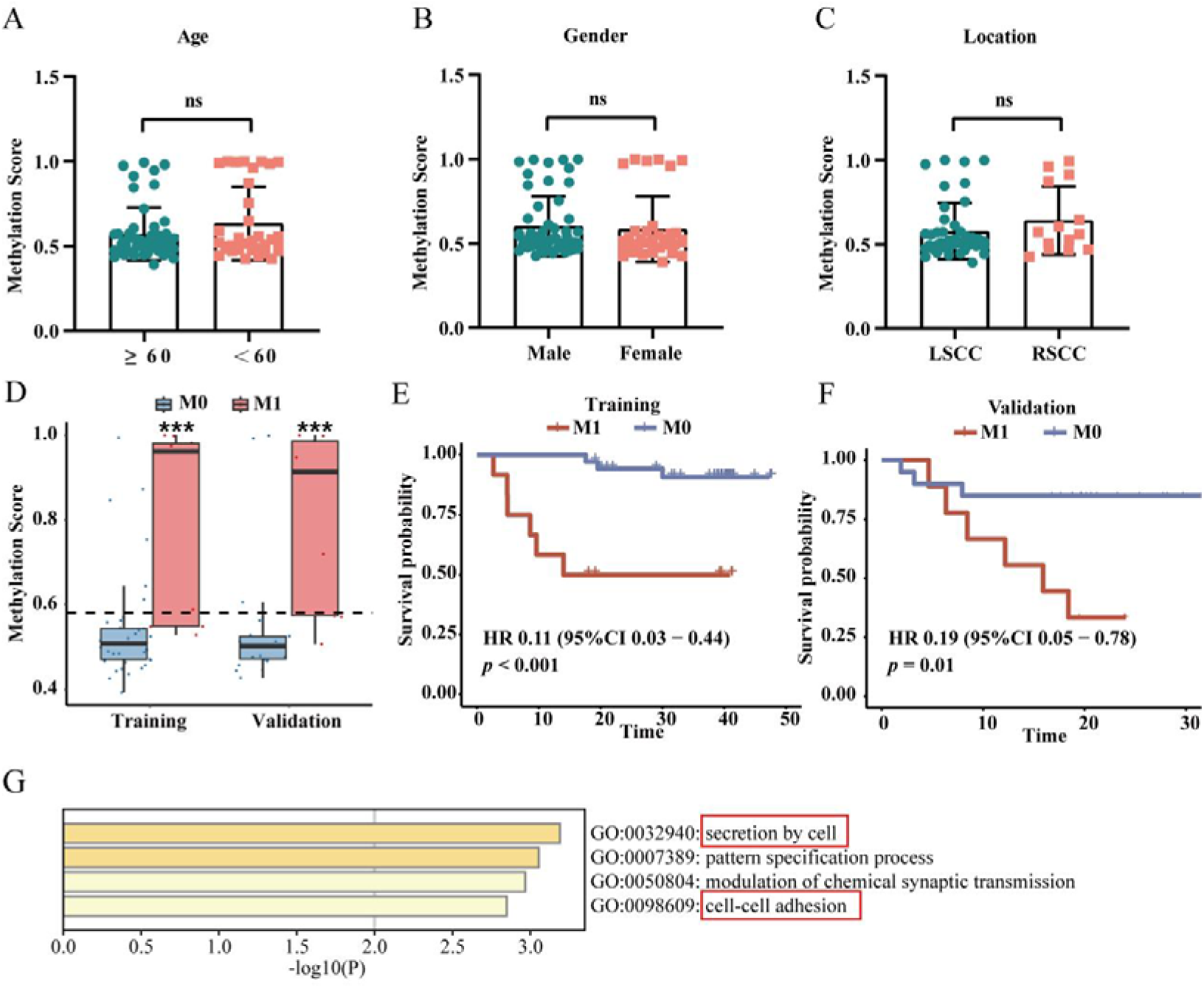
The construction of the metastasis model. (A-C) The methylation scores generated from the diagnosis model for CRC patients with different age (A), gender (B) and location (C). (D) Methylation scores of M0-CRC and M1-CRC individuals generated from the metastasis model (The dashed line represents the cutoff value). (E-F) Kaplan-Meier survival curves comparing OS between the M1-CRC group and M0-CRC group of metastasis model in the training cohort (E) and validation cohort (F). (G) Functional enrichment analysis of genes associated with DMRs exhibiting methylation level changes in blood from patients with AA or CRC 0-II stages.

---

**Table s1.**
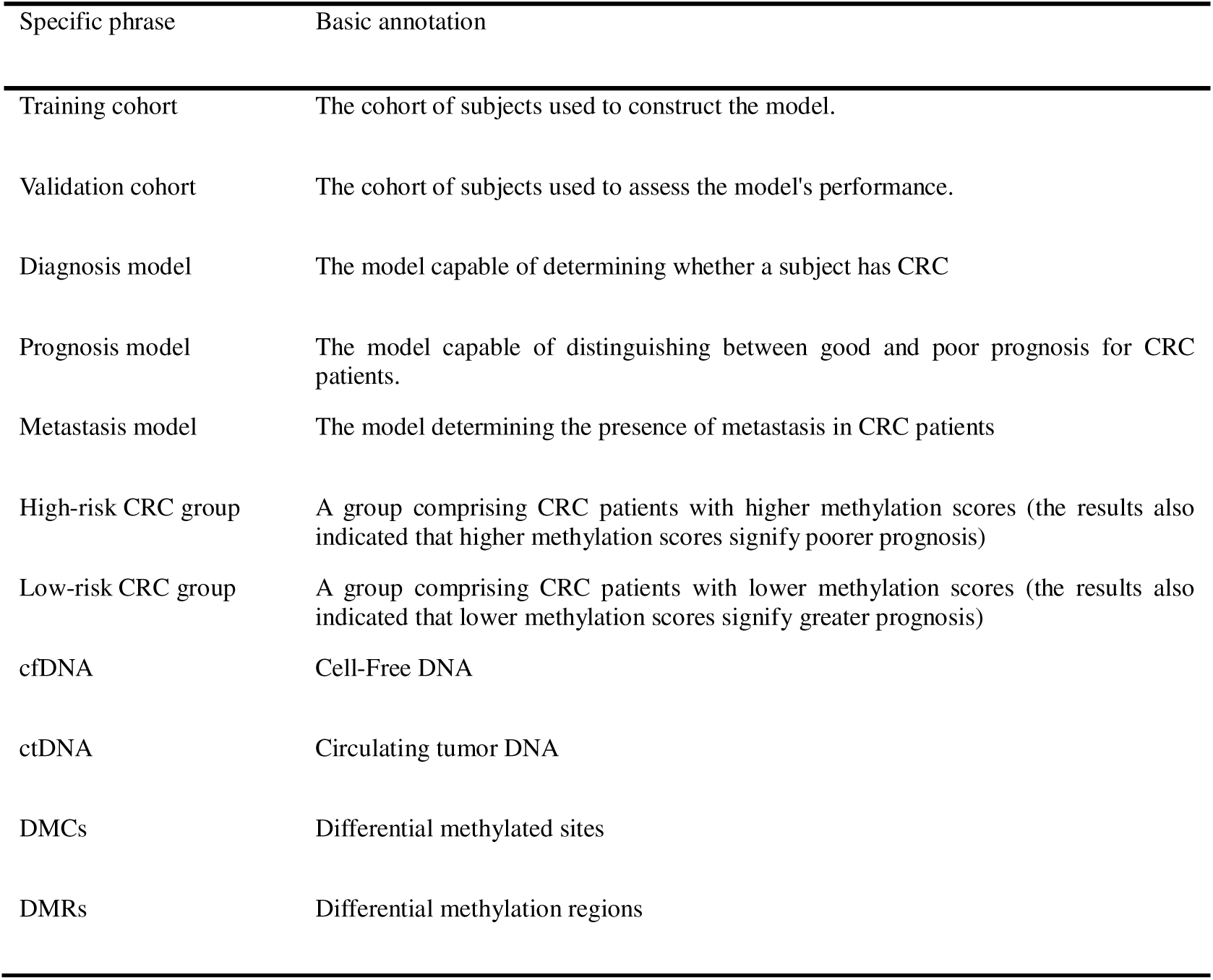
Explanation of specific phrase and abbreviation.

**Table s2.**
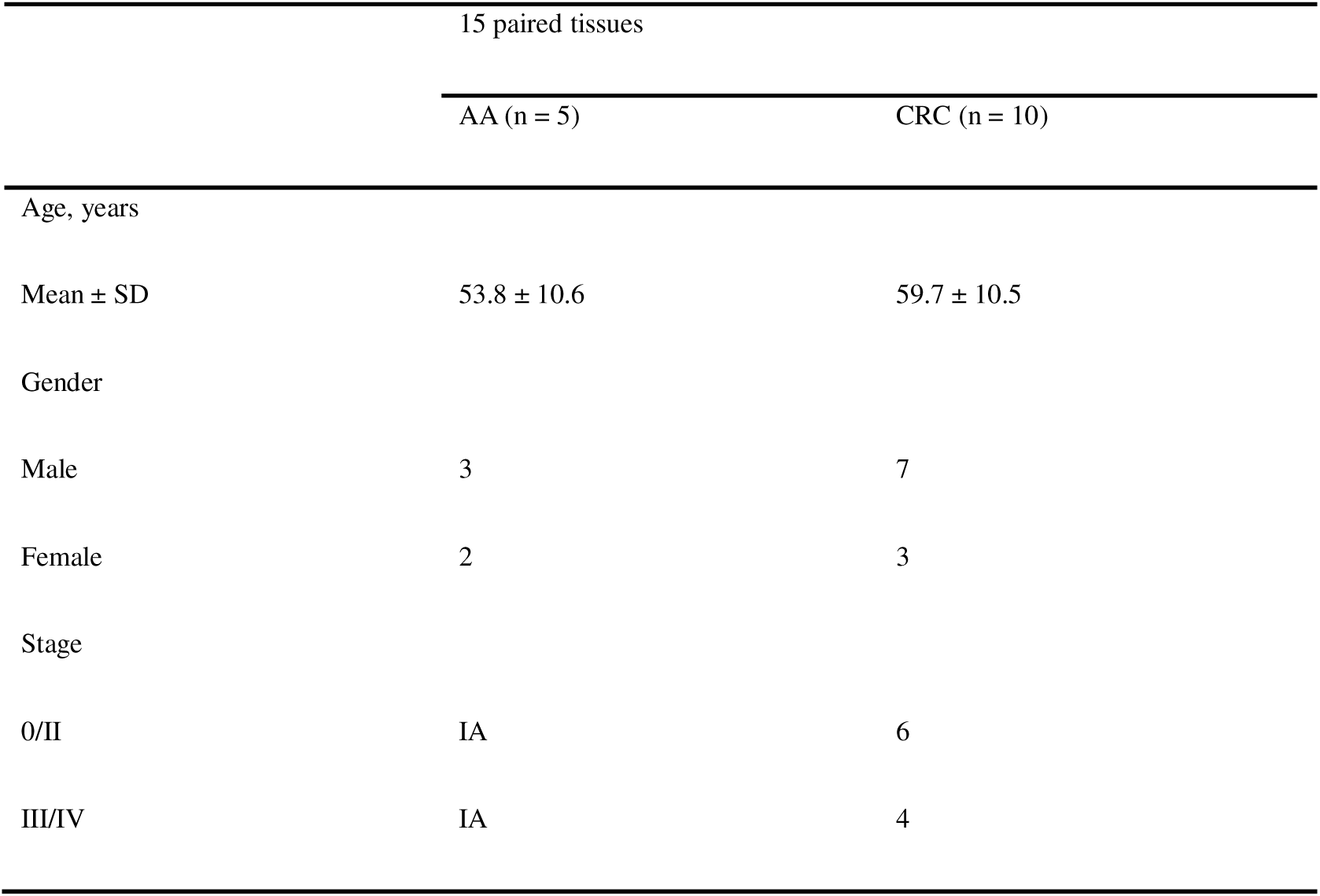
Patient characteristics of the 15 paired tissues.

**Table s3.**
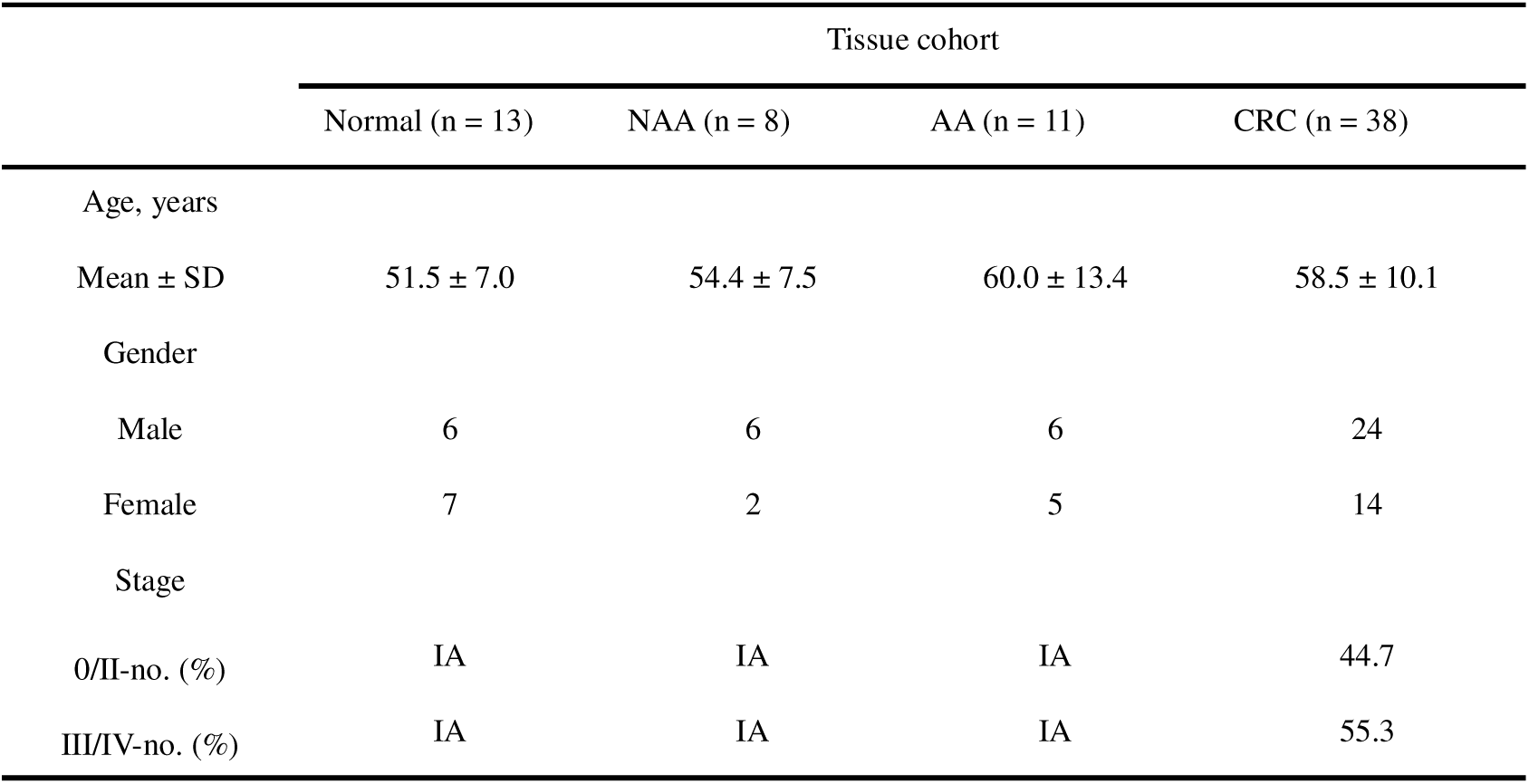
Patient characteristics of the 64 participants (Tissues)

**Table s4.**
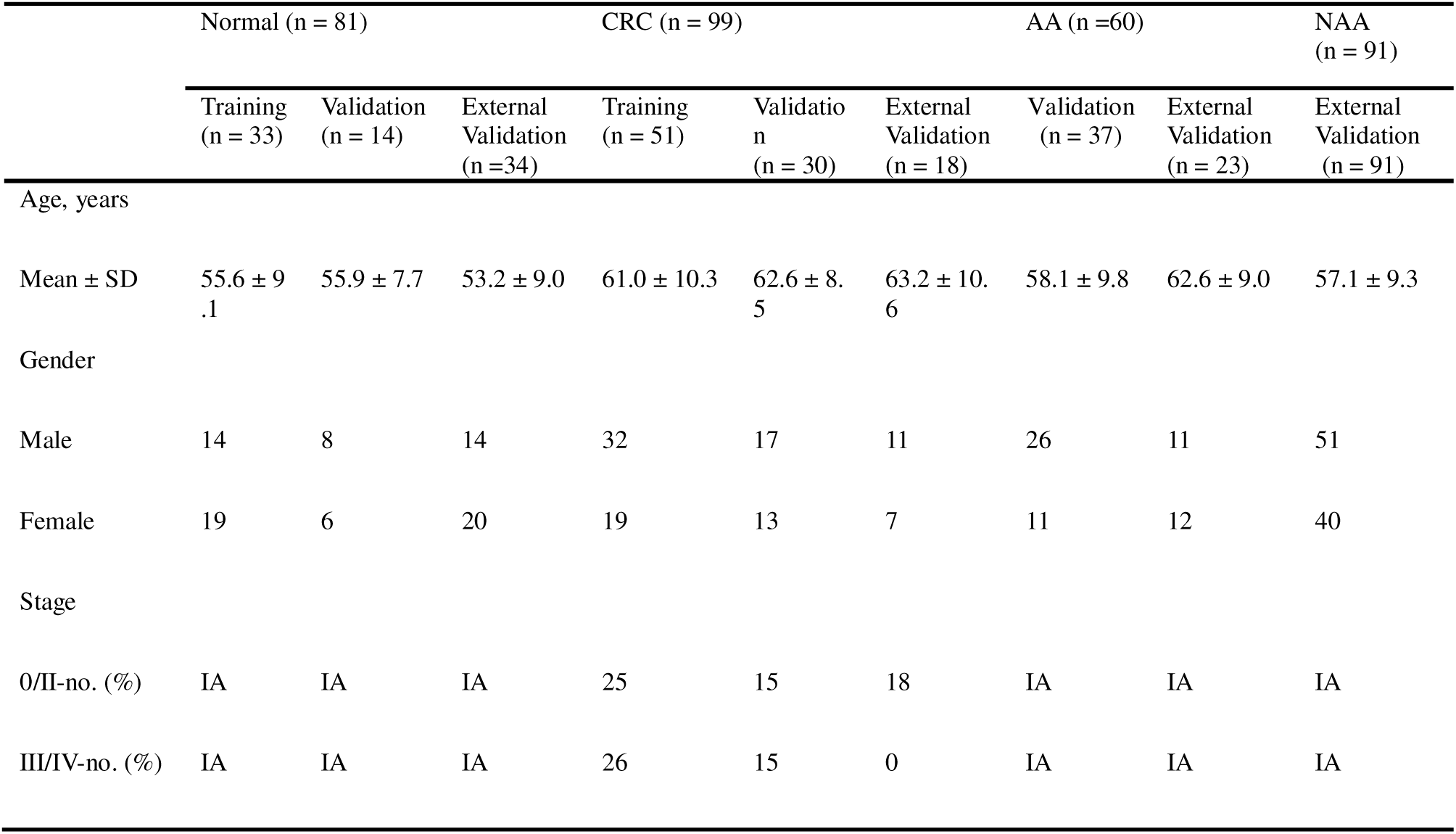
Patient characteristics of the blood cohort.

**Table s5.**
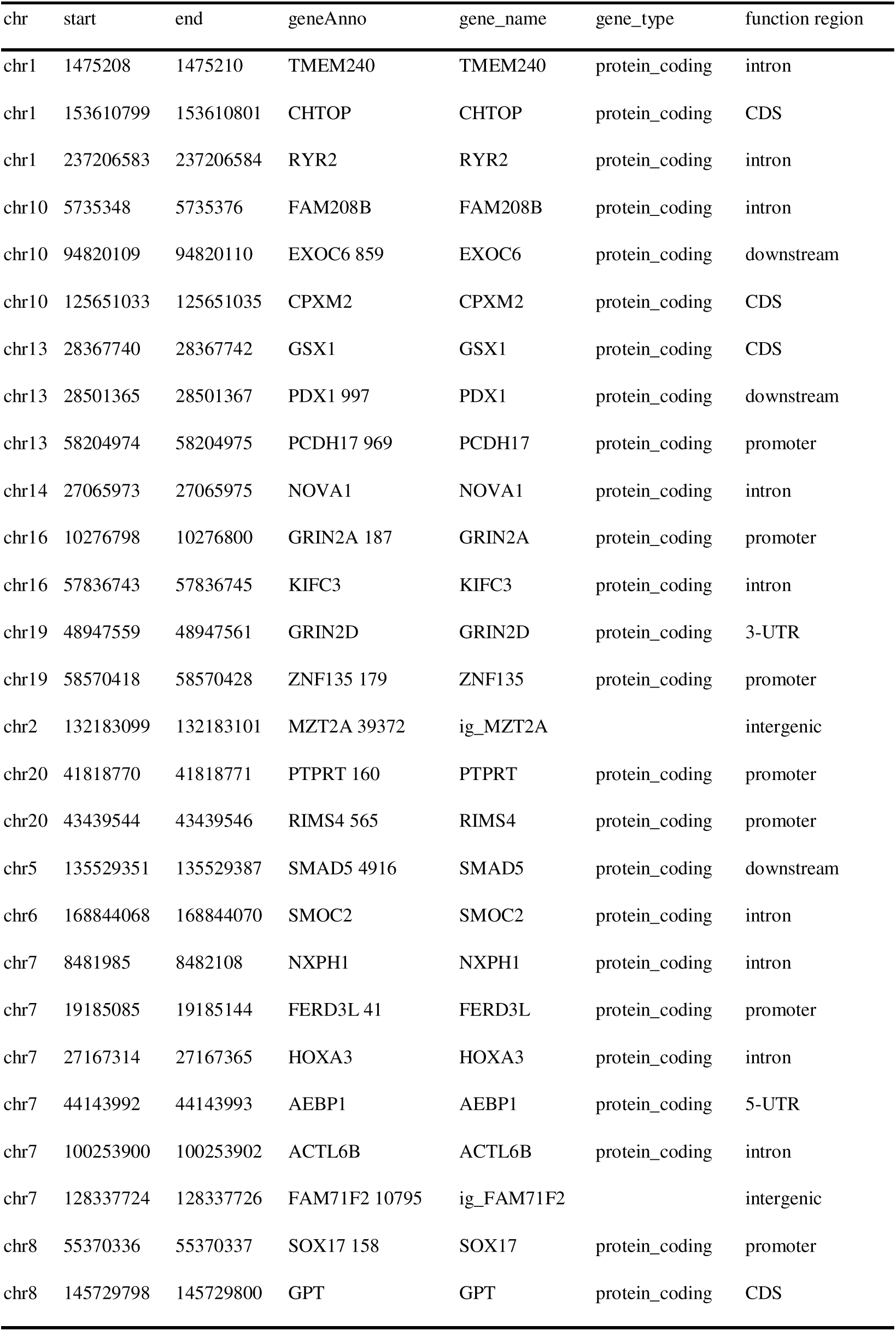
Annotation information for 27 regions.

**Table s6.**
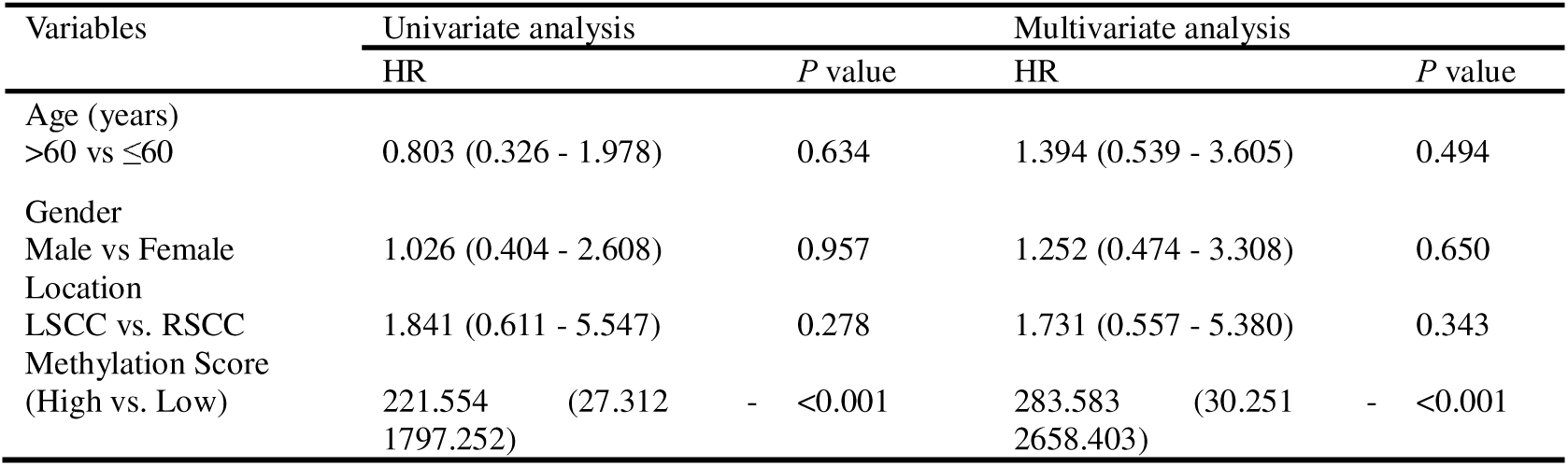
Univariate and multivariate analysis of methylation scores and clinical parameters.

**Table s7.**
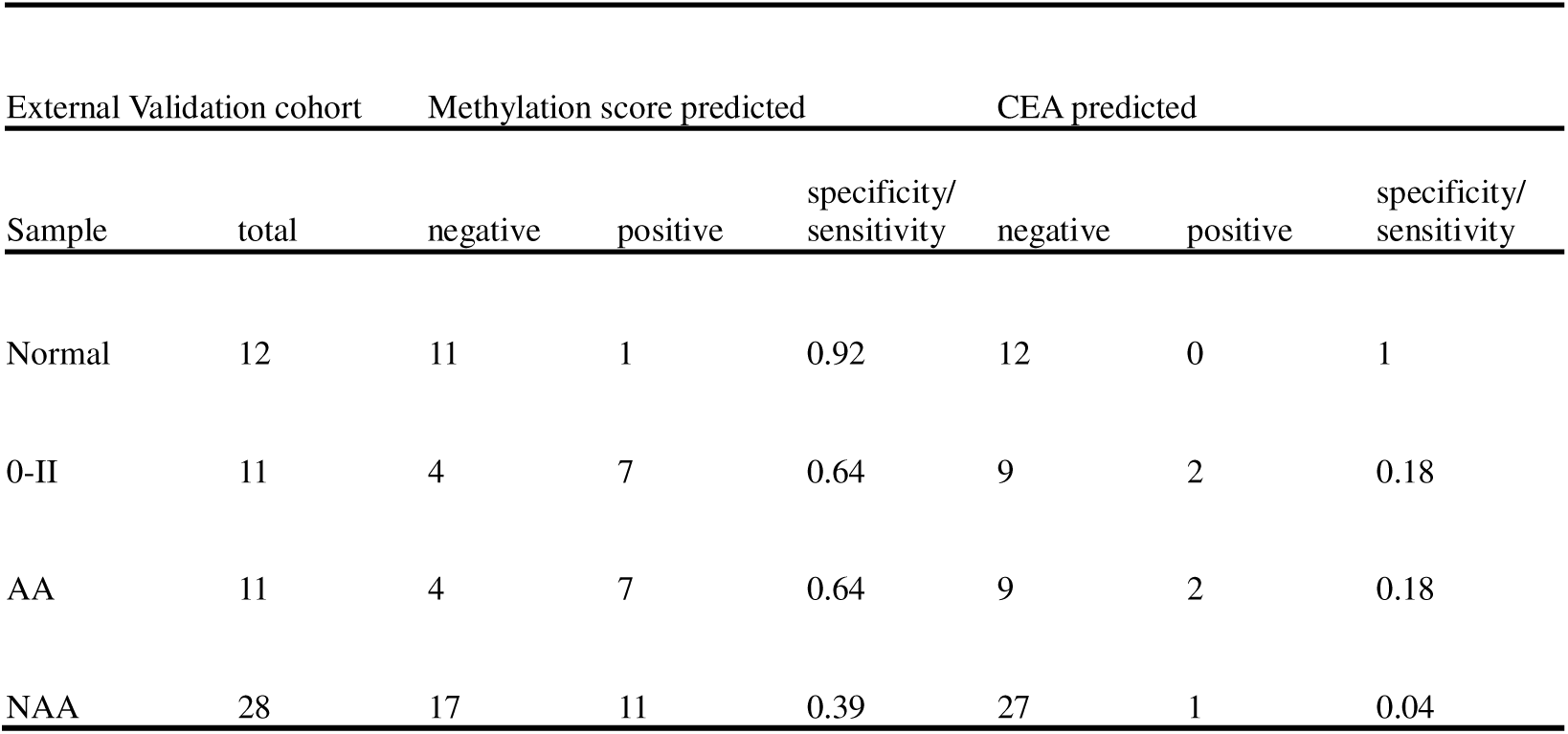
Comparison of diagnostic performance between the methylation score and CEA for early-stage colorectal cancer and adenomas in the external validation cohort.

