## Supplementary material for "Early Diagnosis and Prognostic Prediction of Colorectal Cancer through Plasma Methylation Regions": Supplymentary materials

**Supplementary Figures**

**Supplementary Figure 1**

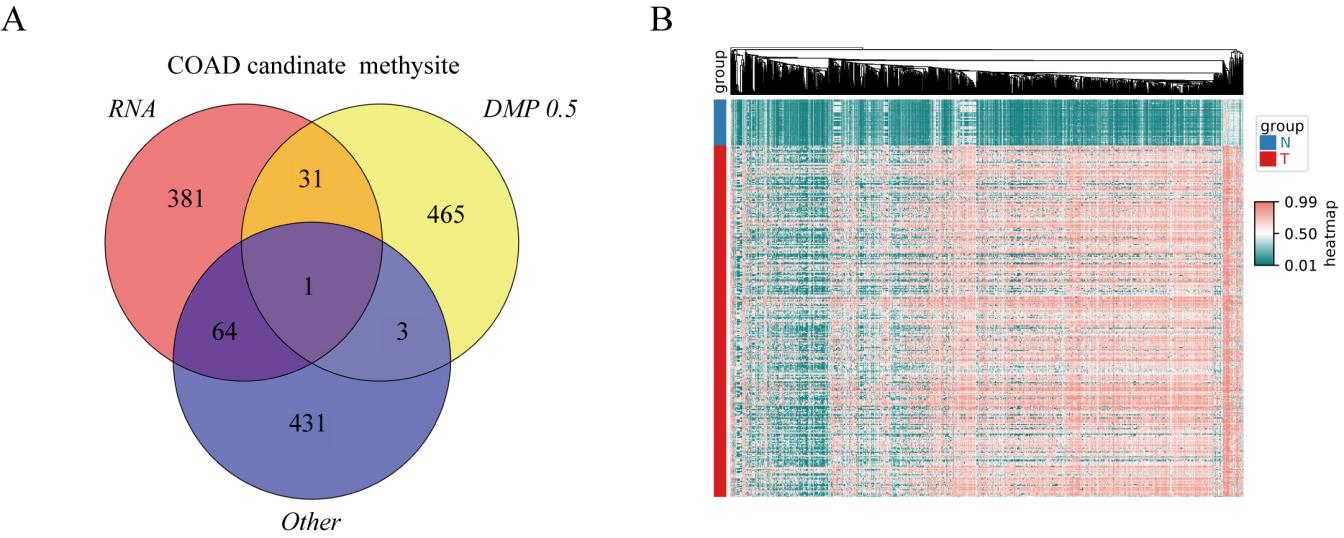

Supplementary Figure 1. Screening for CRC-specific DMCs based on TCGA data. (A) Venn diagram of the results from three DMCs filtering methods based on TCGA datasets. (B) Heatmap illustrating the methylation level of the 1438 methylated sites by unsupervised clustering. Abbreviations: DMCs, differential methylated CpG sites; CRC, colorectal cancer.

**Supplementary Figure 2**

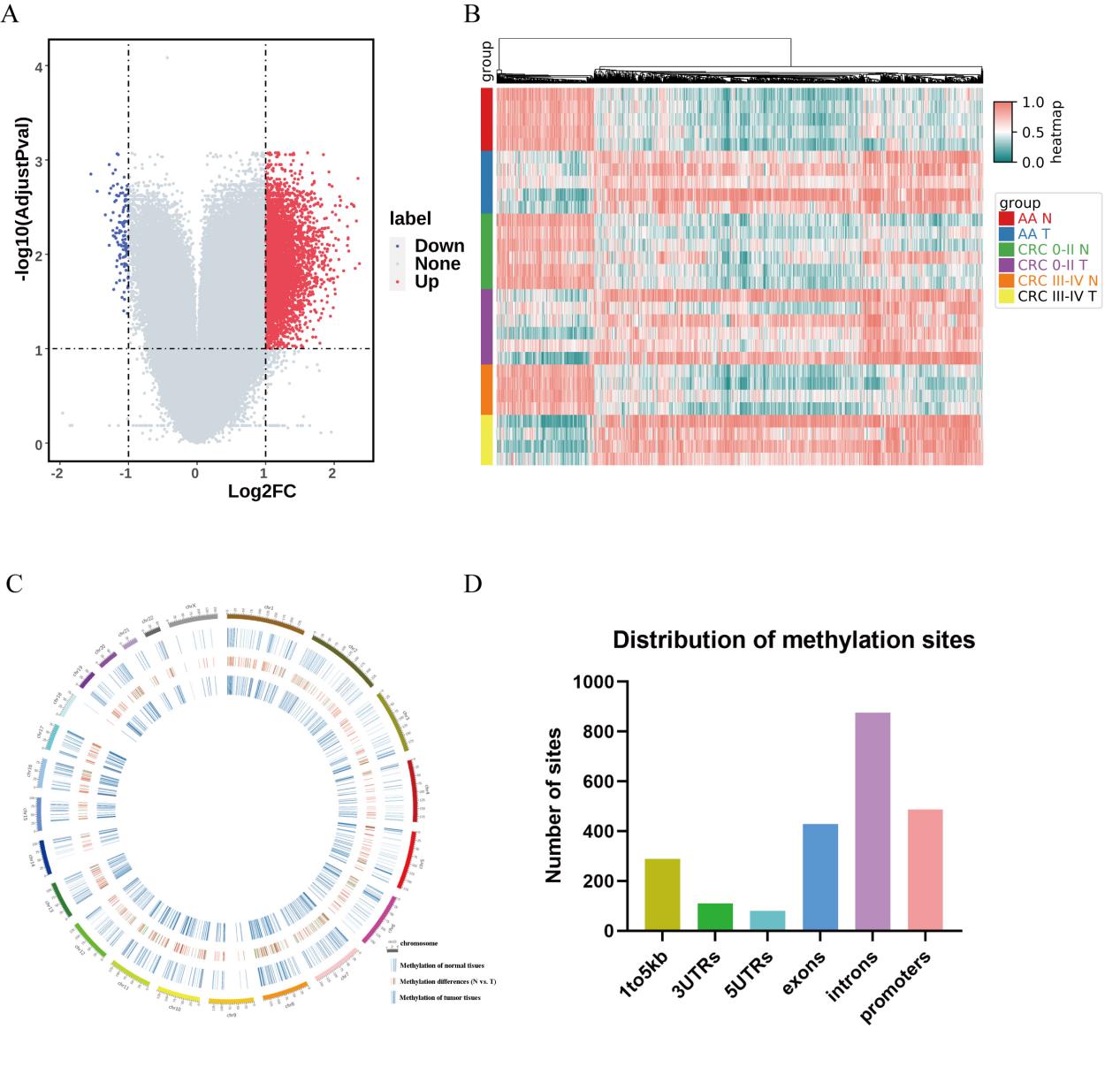

Supplementary Figure 2. DMCs selection in tissues we collected. (A) Volcano plot illustrates the CRC specific DMCs. (B) Heatmap analysis of the top 1400 DMCs between CRC tissues and adjacent tissues by unsupervised clustering. (C) Circular plot of 1400 DMCs depicting methylation level differences between CRC and adjacent tissues. (D) Distribution of 1400 DMCs in gene functional regions. Abbreviations: DMCs, differential methylated CpG sites; CRC, colorectal cancer.

**Supplementary Figure 3**

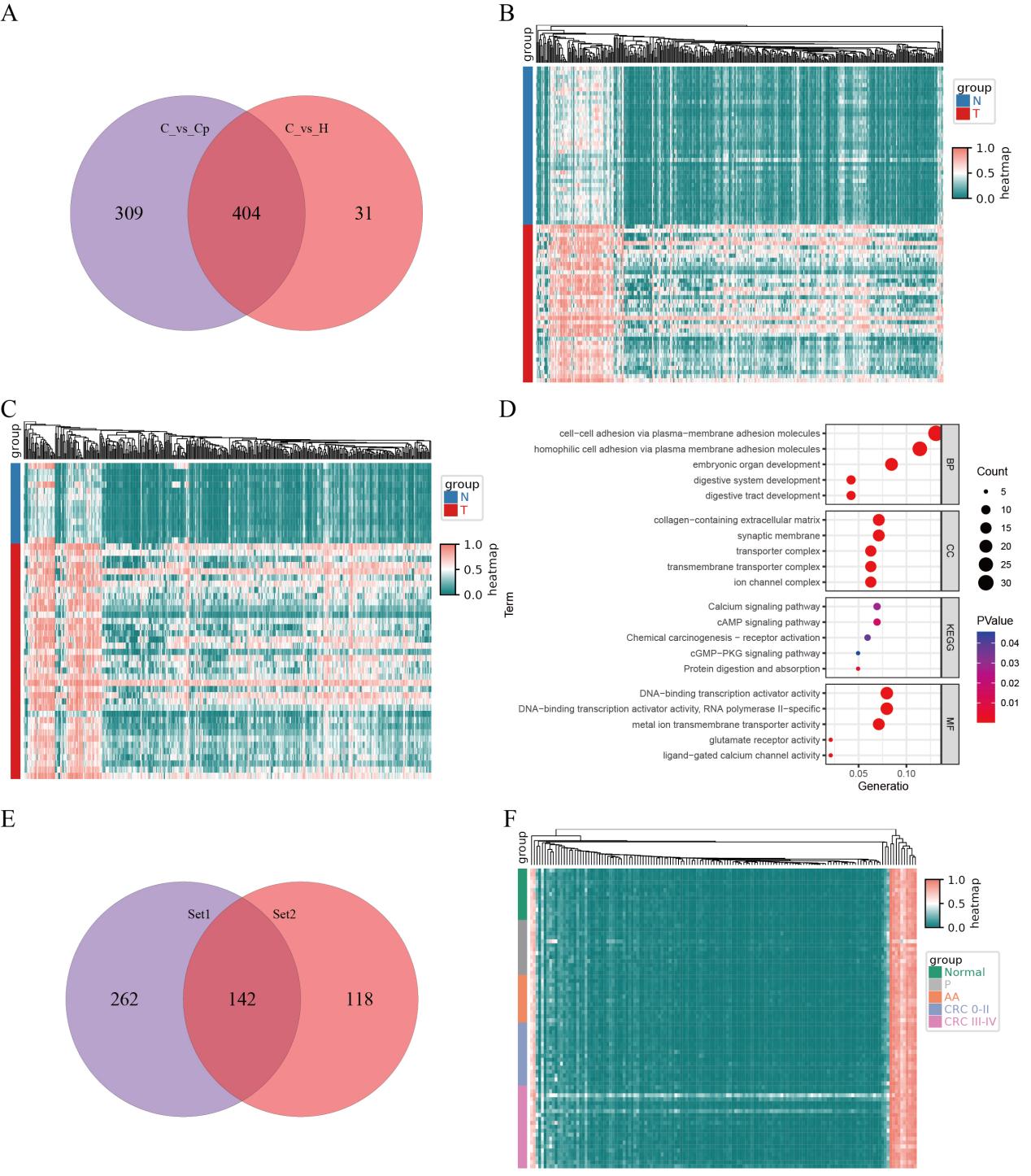

Supplementary Figure 3. Selection of methylation markers. (A) The overlap of DMRs between CRC and adjacent tissues, as well as between CRC and normal tissues. (B) Heatmap illustrating the methylation level of the 404 DMRs in CRC and adjacent tissues. (C) The methylation level of the 404 DMRs in CRC and normal tissues. (D) The gene enrichment analysis for the 404 regions suggests their involvement in processes like cell-cell adhesion, digestive system development, and signaling pathways such as cAMP and cGMP. (E) Based on the results of plasma methylation detection, 142 DMRs were subsequently chosen for further analysis. (F) Heatmap illustrating the methylation levels of the 142 DMRs in the blood samples from normal individuals, those with AA, and patients with CRC. Abbreviations: DMRs, differential methylated regions; AA, adenomatous polyps; CRC, colorectal cancer.

**Supplementary Figure 4**

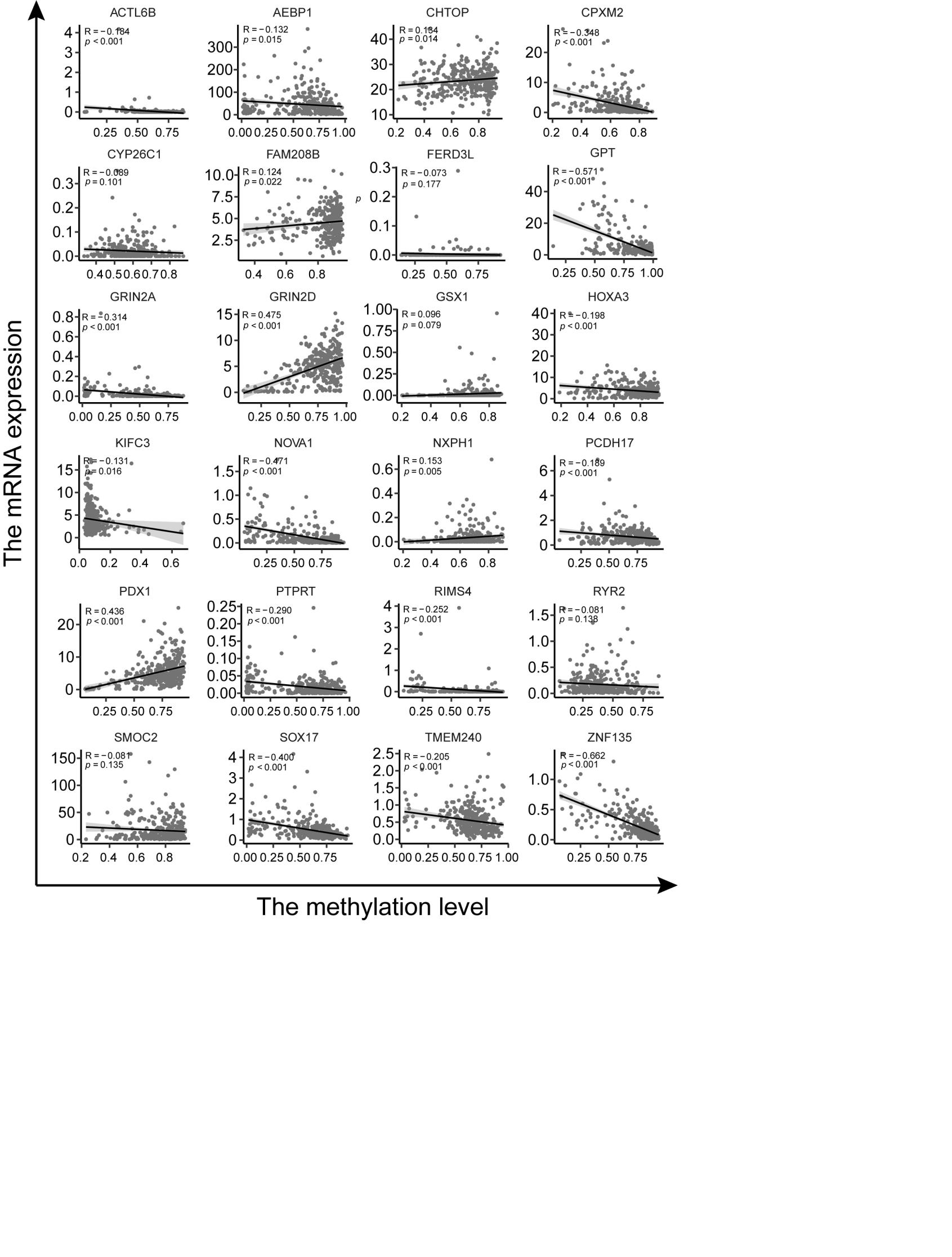

Supplementary Figure 4. The relationship between the methylation levels of the 27 DMRs and the transcription levels of their respective genes in TCGA datasets.

**Supplementary Figure 5**

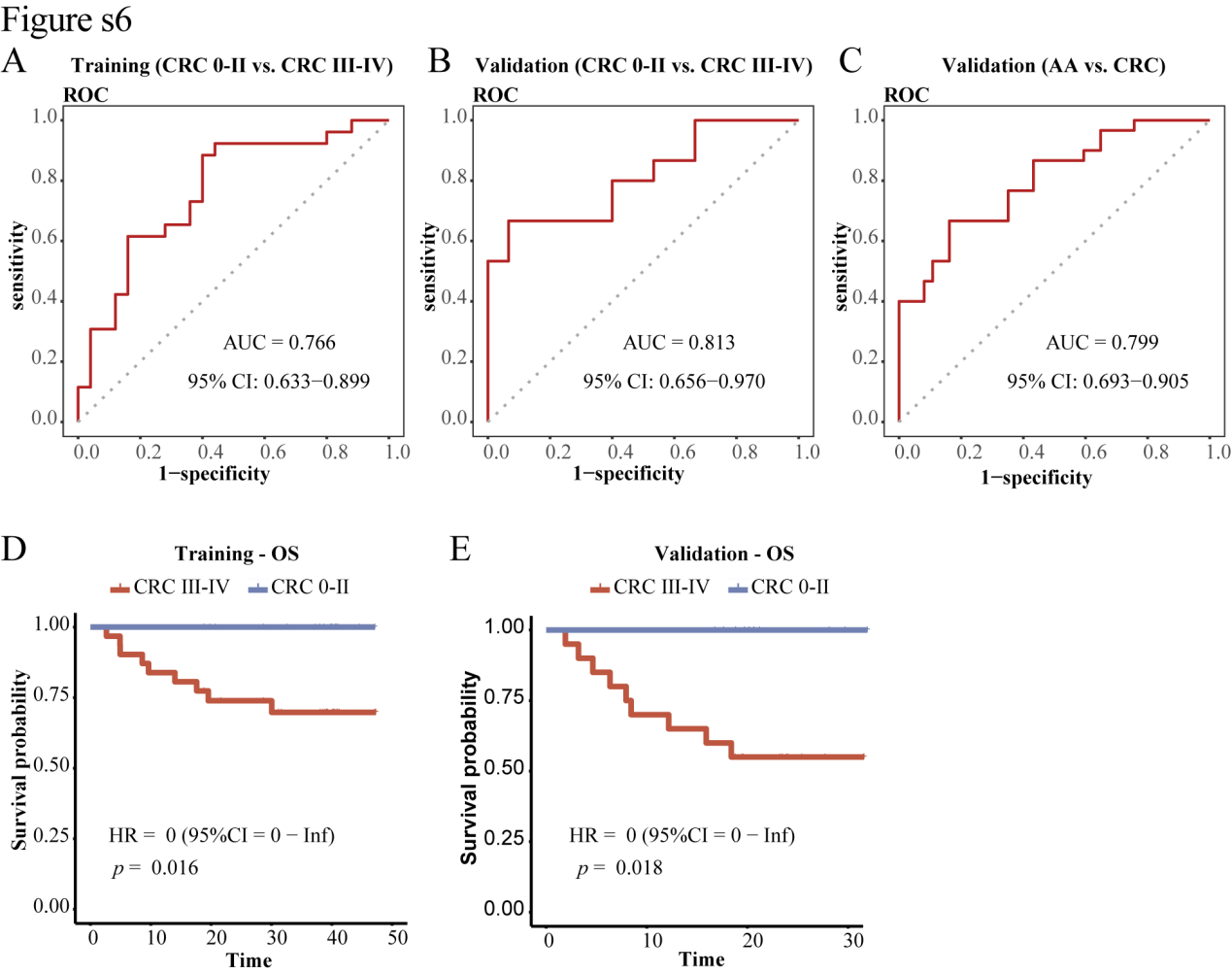

Supplementary Figure 5. Development and validation of a plasma diagnosis model for discerning the specific staging of CRC patients. (A) The use of ROC curve analysis to assess the performance of the diagnostic model in differentiating CRC 0-II patients from CRC III-IV patients of the training cohort. (B-C) The use of ROC curve analysis to evaluate the performance of the diagnostic model in distinguishing CRC 0-II patients from CRC III-IV patients (B), AA from CRC patients (C) of the validation cohort. (D-E) Kaplan-Meier survival curves comparing OS between the CRC III-IV group and CRC 0-II group in the training cohort (D) and validation cohort (E).

**Supplementary Figure 6**

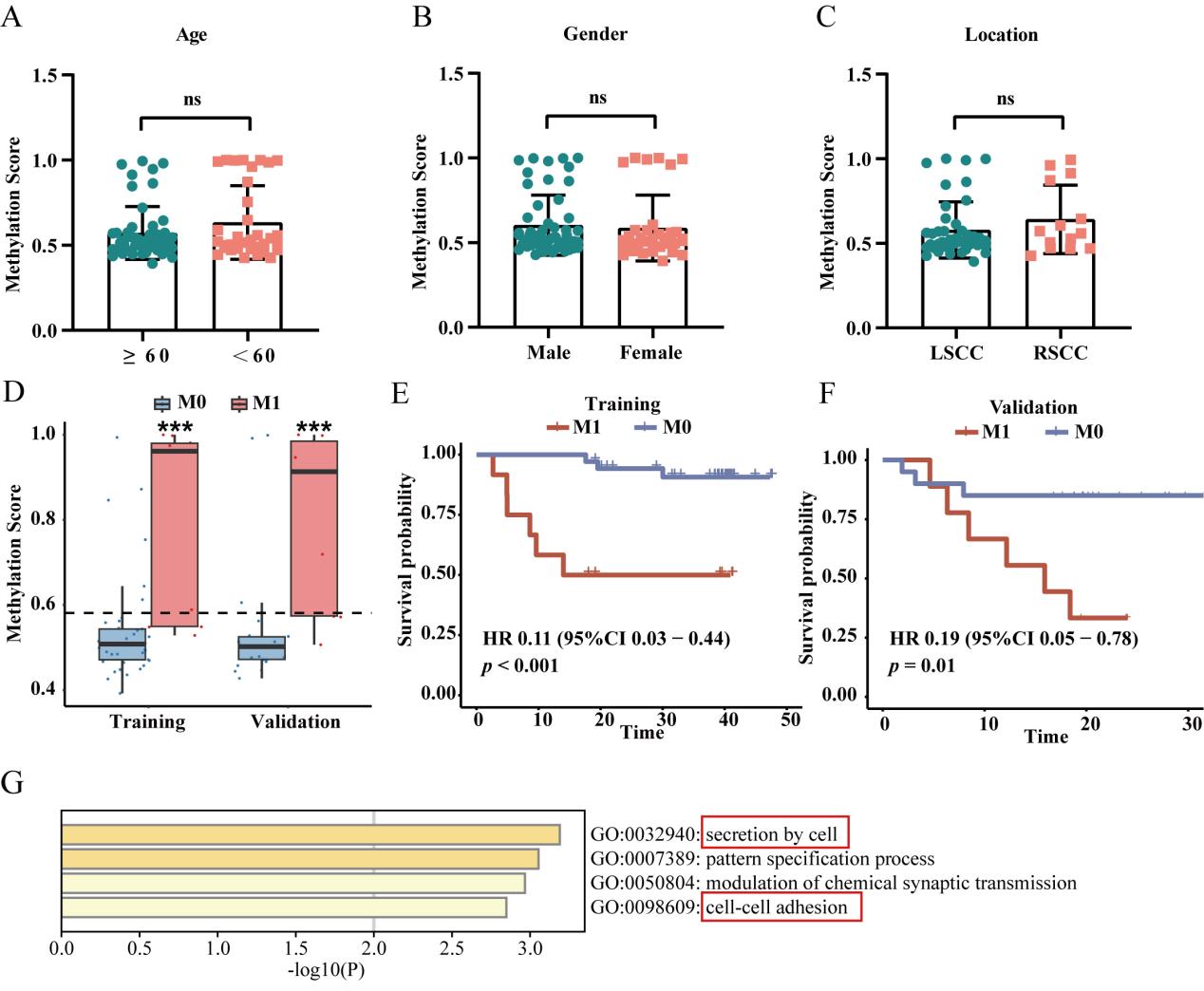

Supplementary Figure 6. The construction of the metastasis model. (A-C) The methylation scores generated from the diagnosis model for CRC patients with different age (A), gender (B) and location (C). (D) Methylation scores of M0-CRC and M1-CRC individuals generated from the metastasis model (The dashed line represents the cutoff value). (E-F) Kaplan-Meier survival curves comparing OS between the M1-CRC group and M0-CRC group of metastasis model in the training cohort (E) and validation cohort (F). (G) Functional enrichment analysis of genes associated with DMRs exhibiting methylation level changes in blood from patients with AA or CRC 0-II stages.

**Supplementary Tables**

**Table s1 Explanation of specific phrase and abbreviation**

| Specific phrase | Basic annotation |
| --- | --- |
| Training cohort | The cohort of subjects used to construct the model. |
| Validation cohort | The cohort of subjects used to assess the model's performance. |
| Diagnosis model | The model capable of determining whether a subject has CRC |
| Prognosis model | The model capable of distinguishing between good and poor prognosis for CRC patients. |
| Metastasis model | The model determining the presence of metastasis in CRC patients |
| High-risk CRC group | A group comprising CRC patients with higher methylation scores (the results also indicated that higher methylation scores signify poorer prognosis) |
| Low-risk CRC group | A group comprising CRC patients with lower methylation scores (the results also indicated that lower methylation scores signify greater prognosis) |
| cfDNA | Cell-Free DNA |
| ctDNA | Circulating tumor DNA |
| DMCs | Differential methylated sites |
| DMRs | Differential methylation regions |

**Table s2 Patient characteristics of the 15 paired tissues**

|  | 15 paired tissues | |
| --- | --- | --- |
|  | AA (n = 5) | CRC (n = 10) |
| Age, years |  |  |
| Mean ± SD | 53.8 ± 10.6 | 59.7 ± 10.5 |
| Gender |  |  |
| Male | 3 | 7 |
| Female | 2 | 3 |
| Stage |  |  |
| 0/II | IA | 6 |
| III/IV | IA | 4 |

**Table s3 Patient characteristics of the 64 participants (Tissues)**

|  | Tissue cohort | | | |
| --- | --- | --- | --- | --- |
|  | Normal (n = 13) | NAA (n = 8) | AA (n = 11) | CRC (n = 38) |
| Age, years |  | | | |
| Mean ± SD | 51.5 ± 7.0 | 54.4 ± 7.5 | 60.0 ± 13.4 | 58.5 ± 10.1 |
| Gender |  | | | |
| Male | 6 | 6 | 6 | 24 |
| Female | 7 | 2 | 5 | 14 |
| Stage |  | | | |
| 0/II-no. (%) | IA | IA | IA | 44.7 |
| III/IV-no. (%) | IA | IA | IA | 55.3 |

**Table s4 Patient characteristics of the blood cohort**

|  | Normal (n = 81) | | | CRC (n = 99) | | | AA (n =60) | | NAA  (n = 91) |
| --- | --- | --- | --- | --- | --- | --- | --- | --- | --- |
|  | Training (n = 33) | Validation (n = 14) | External Validation (n =34) | Training (n = 51) | Validation (n = 30) | External Validation (n = 18) | Validation (n = 37) | External Validation  (n = 23) | External Validation  (n = 91) |
| Age, years |  |  |  |  |  |  |  |  |  |
| Mean ± SD | 55.6 ± 9.1 | 55.9 ± 7.7 | 53.2 ± 9.0 | 61.0 ± 10.3 | 62.6 ± 8.5 | 63.2 ± 10.6 | 58.1 ± 9.8 | 62.6 ± 9.0 | 57.1 ± 9.3 |
| Gender |  |  |  |  |  |  |  |  |  |
| Male | 14 | 8 | 14 | 32 | 17 | 11 | 26 | 11 | 51 |
| Female | 19 | 6 | 20 | 19 | 13 | 7 | 11 | 12 | 40 |
| Stage |  |  |  |  |  |  |  |  |  |
| 0/II-no. (%) | IA | IA | IA | 25 | 15 | 18 | IA | IA | IA |
| III/IV-no. (%) | IA | IA | IA | 26 | 15 | 0 | IA | IA | IA |

**Table s5. Annotation information for 27 regions**

| chr | start | end | geneAnno | gene_name | gene_type | function region |
| --- | --- | --- | --- | --- | --- | --- |
| chr1 | 1475208 | 1475210 | TMEM240 | TMEM240 | protein_coding | intron |
| chr1 | 153610799 | 153610801 | CHTOP | CHTOP | protein_coding | CDS |
| chr1 | 237206583 | 237206584 | RYR2 | RYR2 | protein_coding | intron |
| chr10 | 5735348 | 5735376 | FAM208B | FAM208B | protein_coding | intron |
| chr10 | 94820109 | 94820110 | EXOC6 859 | EXOC6 | protein_coding | downstream |
| chr10 | 125651033 | 125651035 | CPXM2 | CPXM2 | protein_coding | CDS |
| chr13 | 28367740 | 28367742 | GSX1 | GSX1 | protein_coding | CDS |
| chr13 | 28501365 | 28501367 | PDX1 997 | PDX1 | protein_coding | downstream |
| chr13 | 58204974 | 58204975 | PCDH17 969 | PCDH17 | protein_coding | promoter |
| chr14 | 27065973 | 27065975 | NOVA1 | NOVA1 | protein_coding | intron |
| chr16 | 10276798 | 10276800 | GRIN2A 187 | GRIN2A | protein_coding | promoter |
| chr16 | 57836743 | 57836745 | KIFC3 | KIFC3 | protein_coding | intron |
| chr19 | 48947559 | 48947561 | GRIN2D | GRIN2D | protein_coding | 3-UTR |
| chr19 | 58570418 | 58570428 | ZNF135 179 | ZNF135 | protein_coding | promoter |
| chr2 | 132183099 | 132183101 | MZT2A 39372 | ig_MZT2A |  | intergenic |
| chr20 | 41818770 | 41818771 | PTPRT 160 | PTPRT | protein_coding | promoter |
| chr20 | 43439544 | 43439546 | RIMS4 565 | RIMS4 | protein_coding | promoter |
| chr5 | 135529351 | 135529387 | SMAD5 4916 | SMAD5 | protein_coding | downstream |
| chr6 | 168844068 | 168844070 | SMOC2 | SMOC2 | protein_coding | intron |
| chr7 | 8481985 | 8482108 | NXPH1 | NXPH1 | protein_coding | intron |
| chr7 | 19185085 | 19185144 | FERD3L 41 | FERD3L | protein_coding | promoter |
| chr7 | 27167314 | 27167365 | HOXA3 | HOXA3 | protein_coding | intron |
| chr7 | 44143992 | 44143993 | AEBP1 | AEBP1 | protein_coding | 5-UTR |
| chr7 | 100253900 | 100253902 | ACTL6B | ACTL6B | protein_coding | intron |
| chr7 | 128337724 | 128337726 | FAM71F2 10795 | ig_FAM71F2 |  | intergenic |
| chr8 | 55370336 | 55370337 | SOX17 158 | SOX17 | protein_coding | promoter |
| chr8 | 145729798 | 145729800 | GPT | GPT | protein_coding | CDS |

**Table s6 Univariate and multivariate analysis of methylation scores and clinical parameters.**

| Variables | Univariate analysis | | Multivariate analysis | |
| --- | --- | --- | --- | --- |
|  | HR | *P* value | HR | *P* value |
| Age (years) |  |  |  |  |
| >60 vs ≤60 | 0.803 (0.326 - 1.978) | 0.634 | 1.394 (0.539 - 3.605) | 0.494 |
| Gender |  |  |  |  |
| Male vs Female | 1.026 (0.404 - 2.608) | 0.957 | 1.252 (0.474 - 3.308) | 0.650 |
| Location |  |  |  |  |
| LSCC vs. RSCC | 1.841 (0.611 - 5.547) | 0.278 | 1.731 (0.557 - 5.380) | 0.343 |
| Methylation Score |  |  |  |  |
| (High vs. Low) | 221.554 (27.312 - 1797.252) | <0.001 | 283.583 (30.251 - 2658.403) | <0.001 |

**Table s7 Comparison of diagnostic performance between the methylation score and CEA for early-stage colorectal cancer and adenomas in the external validation cohort.**

| External Validation cohort | | Methylation score predicted | | | CEA predicted | | |
| --- | --- | --- | --- | --- | --- | --- | --- |
| Sample | total | negative | positive | specificity/  sensitivity | negative | positive | specificity/  sensitivity |
| Normal | 12 | 11 | 1 | 0.92 | 12 | 0 | 1 |
| 0-II | 11 | 4 | 7 | 0.64 | 9 | 2 | 0.18 |
| AA | 11 | 4 | 7 | 0.64 | 9 | 2 | 0.18 |
| NAA | 28 | 17 | 11 | 0.39 | 27 | 1 | 0.04 |
